# Genomic characterization and molecular evolution of SARS-CoV-2 in Rio Grande do Sul State, Brazil

**DOI:** 10.1101/2023.01.02.23284121

**Authors:** Amanda de Menezes Mayer, Patrícia Aline Gröhs Ferrareze, Luiz Felipe Valter de Oliveira, Tatiana Schäffer Gregianini, Carla Lucia Andretta Moreira Neves, Gabriel Dickin Caldana, Lívia Kmetzsch, Claudia Elizabeth Thompson

**Affiliations:** Center of Biotechnology, Graduate Program in Cell and Molecular Biology (PPGBCM), Universidade Federal do Rio Grande do Sul (UFRGS), Porto Alegre, RS, Brazil; Graduate Program in Health Sciences, Universidade Federal de Ciências da Saúde de Porto Alegre (UFCSPA), Porto Alegre, RS, Brazil; BiomeHub, Florianópolis, Brazil; Laboratório Central de Saúde Pública do Centro Estadual de Vigilância em Saúde da Secretaria de Saúde do Estado do Rio Grande do Sul (LACEN/CEVS/SES-RS), Porto Alegre, RS, Brazil; Department of Pharmacosciences, Universidade Federal de Ciências da Saúde de Porto Alegre (UFCSPA), Porto Alegre, Brazil

**Keywords:** SARS-CoV-2, Genomics, COVID-19, molecular evolution, phylogenomics

## Abstract

The SARS-CoV-2 is the virus responsible for the COVID-19 pandemic and is plaguing the world since the end of 2019. Different lineages have been discovered ever since and the Gamma lineage, which started the second wave of infections, was first described in Brazil, one of the most affected countries by pandemic. Describing the viral genome and how the virus behaves is essential to contain its propagation and to the development of medications and vaccines. Therefore, this study analyzed SARS-CoV-2 sequenced genomes from Esteio city in Rio Grande do Sul, Southern Brazil. We also comparatively analyzed genomes of the two first years of the pandemic from Rio Grande do Sul state for understanding their genomic and evolutionary patterns. The phylogenomic analysis showed monophyletic groups for Alpha, Gamma, Delta and Omicron, as well as for other circulating lineages in the state. Molecular evolutionary analysis identified several sites under adaptive selection in membrane and nucleocapsid proteins which could be related to a prevalent stabilizing effect on membrane protein structure, as well as majoritarily destabilizing effects on C-terminal nucleocapsid domain.

## 1 INTRODUCTION

After the first outbreak of COVID-19 (Coronavirus disease 2019) in Wuhan, Hubei Province, China in December 2019 (Zhu et al., 2020), the new severe acute respiratory syndrome coronavirus 2 (SARS-CoV-2) spread around the world. Starting a world pandemic, declared by the World Health Organization (WHO) in March 2020, COVID-19 already exceeds 649 million cases and 6.65 million deaths until December, 2022 (Dong et al., 2020, https://coronavirus.jhu.edu/map.html accessed on December 13, 2022). Currently (December, 2022), Brazil is the fifth country most affected by SARS-CoV-2, reaching the mark of 35 million confirmed cases and more than 690 thousands deaths (Dong et al., 2020, https://coronavirus.jhu.edu/map.html accessed on December 13, 2022). Of these, 7.9% of the cases and 6% of the deaths are from Rio Grande do Sul (RS), the southernmost state of Brazilian territory and the fourth state in the ranking of COVID-19 cases (https://covid.saude.gov.br/, accessed on December 13, 2022).

Among several lineages, along these two years of pandemic, various Variants of Concern (VOCs), such as Alpha, Beta, Gamma, Delta, and Omicron, carrying signature aminoacid substitutions (especially in the spike protein) circulated in RS state (Gularte et al., 2022; Wink et al., 2022; Gräf et al., 2022). The sublineage P.1.2, a Gamma-like variant also considered as a Variant of Interest, was initially identified in RS state (Franceschi et al., 2021), despite other studies estimating its divergence between late 2020 and early 2021 in São Paulo state (Junior et al., 2021). As in Brazil, an increasing number of cases and deaths by Gamma (P.1 lineage) became evident in RS in the beginning of 2021, causing a second COVID-19 wave. The P.1 lineage harbors mutations in the Spike’s receptor-binding domain (RBD) such as E484K, K417T, and N501Y, which promote evasion from antibody neutralization elicited by infection or vaccination (R. E. Chen et al., 2021; Chakraborty, 2022).

In December, 2020, the Delta variant was described initially in India. This variant is highly transmissible and spreads easily, causing new waves of infection around the world by the middle 2021 (Shiehzadegan et al., 2021). However, despite this lineage rapidly becoming dominant in Brazil (including RS state), in July/August, 2020, there was not a concurrent increase in reported cases or deaths (Giovanetti et al., 2022). By the end of 2021, the number of cases of COVID-19 were progressively decreasing, until the emergence of the Omicron variant, confirmed in November 2021, in South Africa (Wang & Powell, 2021). This new variant is a concern due to the mutations on RBD and cleavage sites that suggest higher transmissibility, up to three times more contagious than Delta variant (J. Chen et al., 2022).

Despite all VOC characterizations mostly focused on spike mutations, structural proteins E (Envelope), M (Membrane), and N (Nucleocapsid) are functionally important to the virus assembly and pathogenesis (Yadav et al., 2021). The N protein has been associated to the promotion of inflammatory processes by activation of cyclooxygenase-2 (COX-2), to interaction with p42 proteasome in order to avoid the degradation of viral proteins, and to inhibition of IFN-I in immune response (Satarker & Nampoothiri, 2020). Moreover, M protein is known to inhibit NFκB, to reduce levels of COX-2, to activate IFN-β, and to interact with PDK1/PKB proteins, leading to cell death or apoptosis.

In this way, this study aims to perform genomic sequencing and characterization of the SARS-CoV-2 genomes from RS state as well as to identify selection traits in E, M, and N protein sites, elucidating the molecular evolution processes that drive the diversification or conservation of the structural proteins from SARS-CoV-2.

## 2 RESULTS

### 2.1 Sample Characterization

Twelve respiratory secretion samples used for COVID-19 diagnostic purposes (RT-qPCR) were collected from patients from the municipality of Esteio, Rio Grande do Sul (RS) state, between April 9th and June 29th, 2021. The mean cycle threshold (Ct) value for the first RT-qPCR conducted at Laboratório Central de Saúde Pública do Estado do Rio Grande do Sul (LACEN) was 23.83 (median: 23.00; IQR: 4.5).

The sequence coverage for the twelve sequenced genomes ranged between 84.92 and 99.76% (mean: 98.26%) of the 29,903 bp of NC_045512.2 reference genome. The mean of sequencing depth was calculated to 292.23x, with a variation between 53.15 and 542.28x. Leastwise 48.81% of the sequence accomplished a coverage depth ≥51x (max: 98.27%, mean: 87.68%) (Supplementary File 1). According to Pango lineage assignment, 10 sequenced samples belong to P.1 lineage and 2 are from P.1.17 sublineage, both from Gamma variant clade.

### 2.2 Comparative Genomics of Esteio SARS-CoV-2 sequences

Sixty-eight different nucleotide substitutions were found in the twelve genomes from this study, being 36 of these non-synonymous. The mutations NSP13:E341D (ORF1b:E1264D), S:D614G and S:V1176F were predominantly found in these sequences. The most common missense substitutions (in at least 75% of the sequences) were NSP12:P323L (ORF1b:P314L), S:K417T, S:T1027I, ORF8:E92K, and N:P80R (in 11 genomes); 5’UTR:C241T and NSP3:K977Q (10 genomes) and H655Y (in 9 genomes). Only two samples carry substitutions S:E484K and S:N501Y, being both mutations in the same samples. The mutations S:E661D and S:S689I were present in one sample each.

A few mutations found in our samples are described for the first time in RS (Table 1). Most of them were already identified in the world and occurred in sequences from different VOCs such as Alpha, Beta, Gamma, Delta, and Omicron. Substitution D1208A (NSP3) was also found for the first time in Brazil. The non-synonymous mutation I136V, in the NSP12 gene, was not described on GISAID, being the first report about this mutation (Table 1).

**TABLE 1.** List of mutations firstly described in Rio Grande do Sul in the sequenced genomes from this study.

| Mutation | Occurrences<br>in the world | First<br>occurrence<br>in world | Occurrences<br>in Brazil | First<br>occurrence<br>in Brazil | Date of our<br>sample | VOCs |
| --- | --- | --- | --- | --- | --- | --- |
| ORF3a:<br>A72S | 7,462 | 2020 | 85 | 2021-02-03 | 2021-05-01 | A, B, G,<br>D, O |
| NSP3:<br>P1103L | 17,219 | 2020 | 428 | 2020-06-16 | 2021-06-14 | A, B, G,<br>D, O |
| NSP3:<br>D1208A | 271 | 2020-03-13 | not described | - | 2021-06-14 | A, G, D,<br>O |
| NSP4:<br>D144G | 198 | 2020 | 2 | 2021-05-31 | 2021-06-14 | A, G, D,<br>O |
| NSP13:<br>L325F | 1,255 | 2020 | 15 | 2021-01-27 | 2021-06-14 | A, B, G,<br>D, O |
| NSP12:<br>I136V | not described | - | not described | - | 2021-05-08 | G |

The table describes the mutations and the number of occurrences in the world and Brazil, as well as their first occurrences in these locations. The table also indicates the VOC lineages where each mutation can be found (Access on GISAID: June 7, 2022). A: Alpha; B: Beta; G: Gamma; D: Delta; O: Omicron.

### 2.3 Comparative Genomics of Rio Grande do Sul SARS-CoV-2 sequences

A number of 4,706 sequences were downloaded from the GISAID platform ranging from March, 2020 up to May, 2022 (26 months) (Submission up to September 30, 2022). The genome sequencing count per month in RS state can be visualized in Figure 1. Most sequencing efforts are associated with COVID-19 waves caused by the introduction of new lineages in RS state. This can be observed during Gamma / Delta waves (February up to October, 2021) and, recently, the Omicron wave, started in January, 2022. The total number of sequences in the state is low compared to the number of cases, representing around 0.19% of them (number of cases = 2,435,883 on May 31, 2022).

**FIGURE 1.**
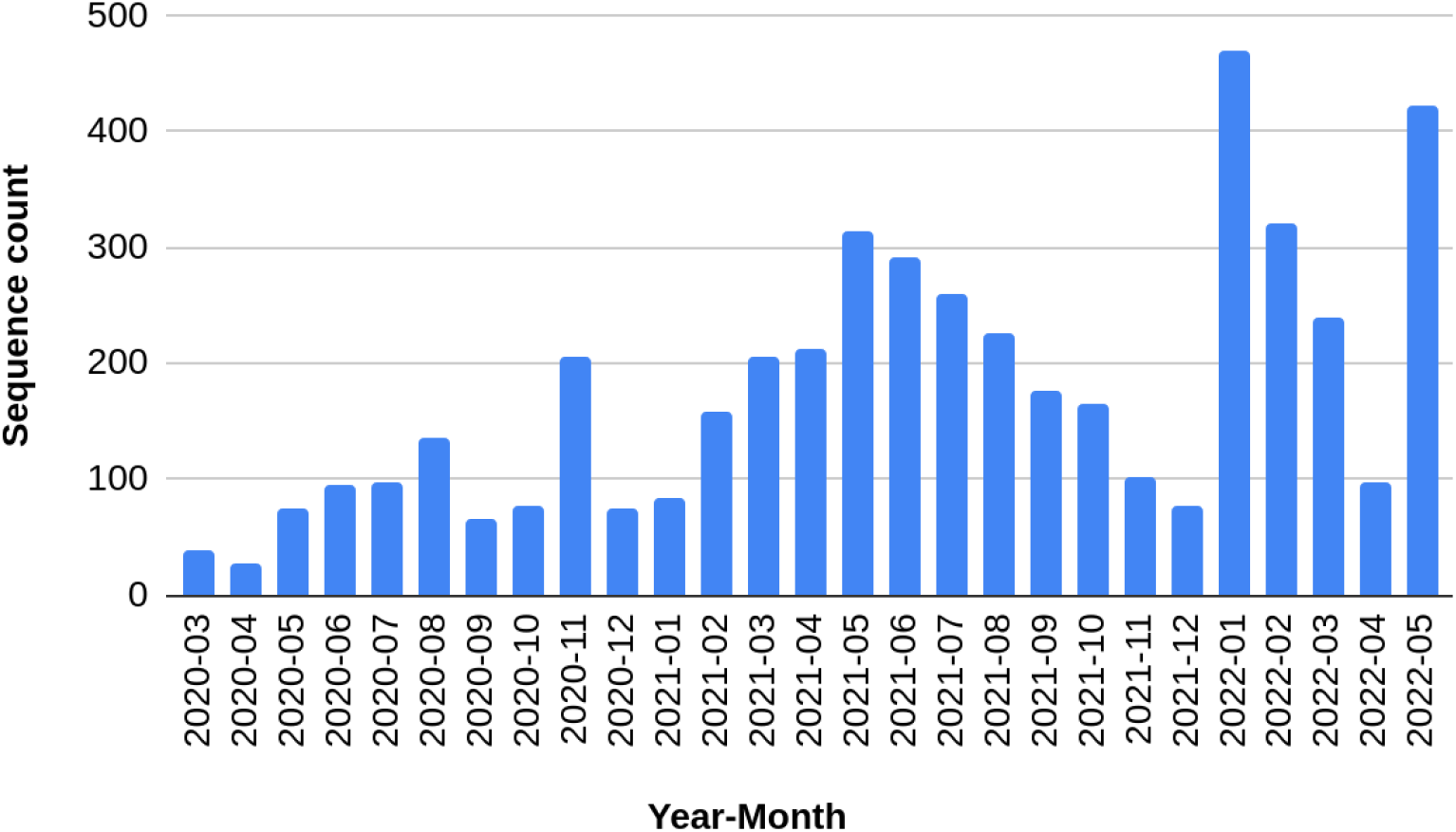
Counts of sequences per month in Rio Grande do Sul between March 2020 and May 2022.

As shown in Figure 2, lineage B.1.1.33 predominates in 2020. Between November 2020 and January 2021, the Zeta (P.2) lineage became more frequent, followed by B.1.1.28 and P.7 lineages. From February until July 2021, P.1 and derivative lineages (mostly P.1 and P.1.2) became prevalent in the RS genomes, in accordance with the lineages of our samples, which were collected between April and June 2021, during the Gamma-related second wave of COVID-19 in the state. Approximately 27.6% of these SARS-CoV-2 genomes obtained on the GISAID database between March 2020 and May 2022 belong to the Gamma clade. Considering only 2021, 57.1% of the SARS-CoV-2 sequenced genomes were classified as Gamma. The Delta lineages (mostly AY.99.2 and AY.101) were initially identified in the state by sequencing in June 2021 and became prevalent from August up to the end of the year. Delta lineages were accountable for 32.30% of the sequenced genomes from RS state in 2021. The Omicron lineages (mostly BA.1.1 and BA.2) arised in RS state in December, 2021, achieving more than 90% of sequenced genomes between January and April, 2022.

**FIGURE 2.**
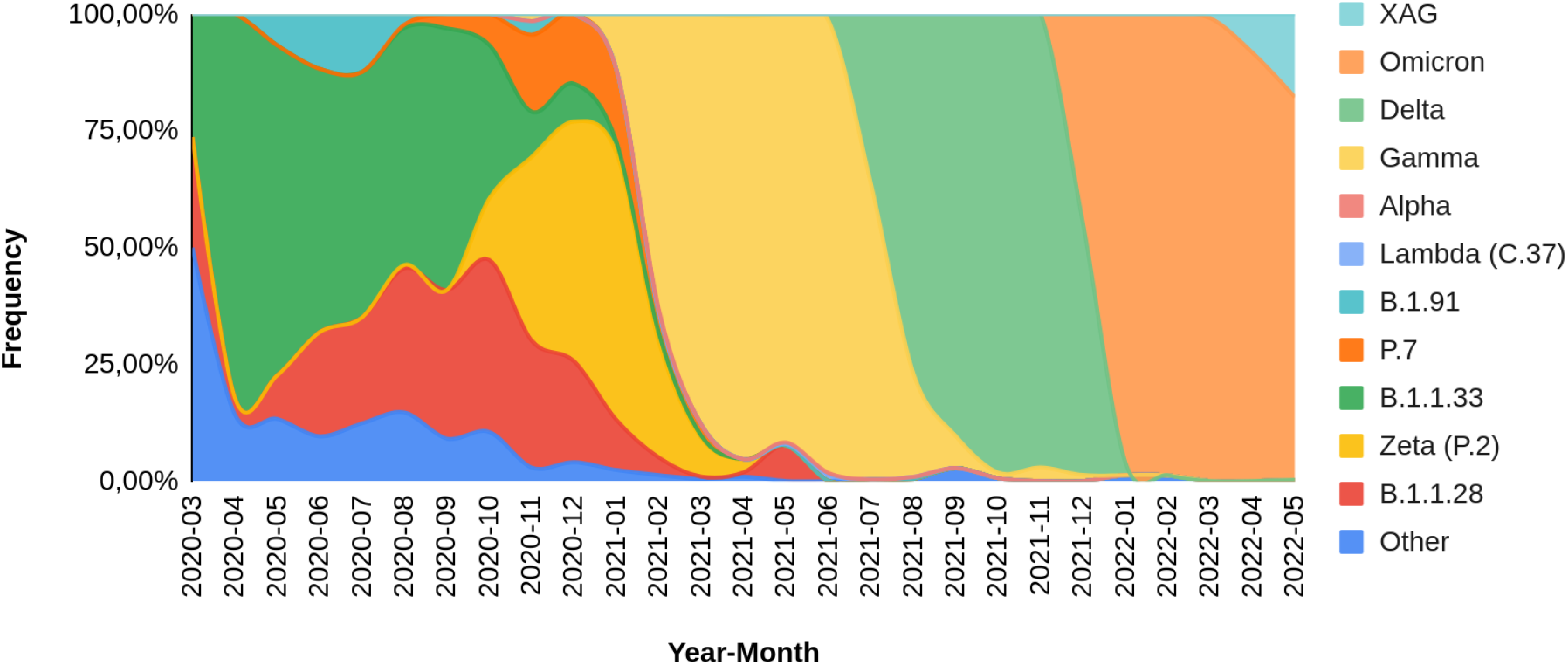
Lineage distribution in Rio Grande do Sul state between March 2020 and May 2022.

About the genome set from RS state, the missense variant NSP12:P323L (ORF1b:P314L) was the most prevalent, found in 98.70% (n = 4,645) of samples (Figure 3). The highly frequent mutations (present in at least 90% of the genomes) also include the extragenic substitution C241T (n = 4,484), the synonymous mutation NSP3:F106F (n = 4,509), and the non-synonymous mutation S:D614G (n = 4,504). Other non-synonymous mutations such as N:R203K/G204R (n = 3,856/3,854), S:N501Y (n = 2,762), S:H655Y (n = 2,842) were found in > 50% of samples.

**FIGURE 3.**
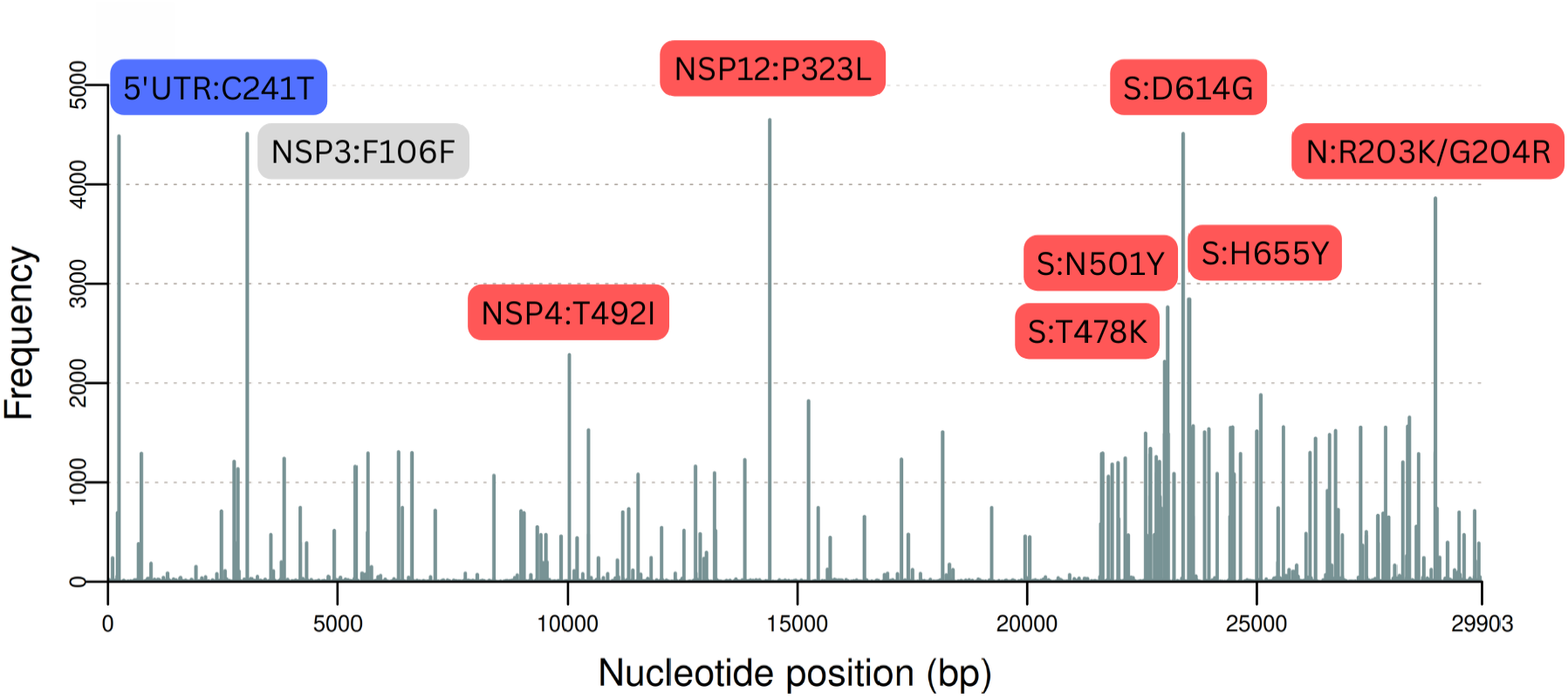
Amino acid and nucleotide substitutions associated with SARS-CoV-2 genomes from RS state. Synonymous and non-synonymous mutations are labeled with the amino acid residue. Substitutions in extragenic positions were labeled with the nucleotide alteration. Mutations occurring in more than 40% of the samples were labeled in different colors: red (non-synonymous mutations), gray (synonymous mutations), and blue (extragenic mutations).

### 2.4 Global Phylogenomics

In order to establish the evolutionary relationships of Esteio sequenced genomes with SARS-CoV-2 global dataset, the AudacityInstant tool from GISAID database was used to identify genetically related genomes. Four sequences could not be related to other sequences in the database, probably due to low sequencing quality. Considering the most related genomes (Table 2), four sequences were associated with samples from Brazil (one of them from Rio Grande do Sul state) and the other four were more closely related to genomes from Chile, Mexico, USA, and Canada.

**TABLE 2.**
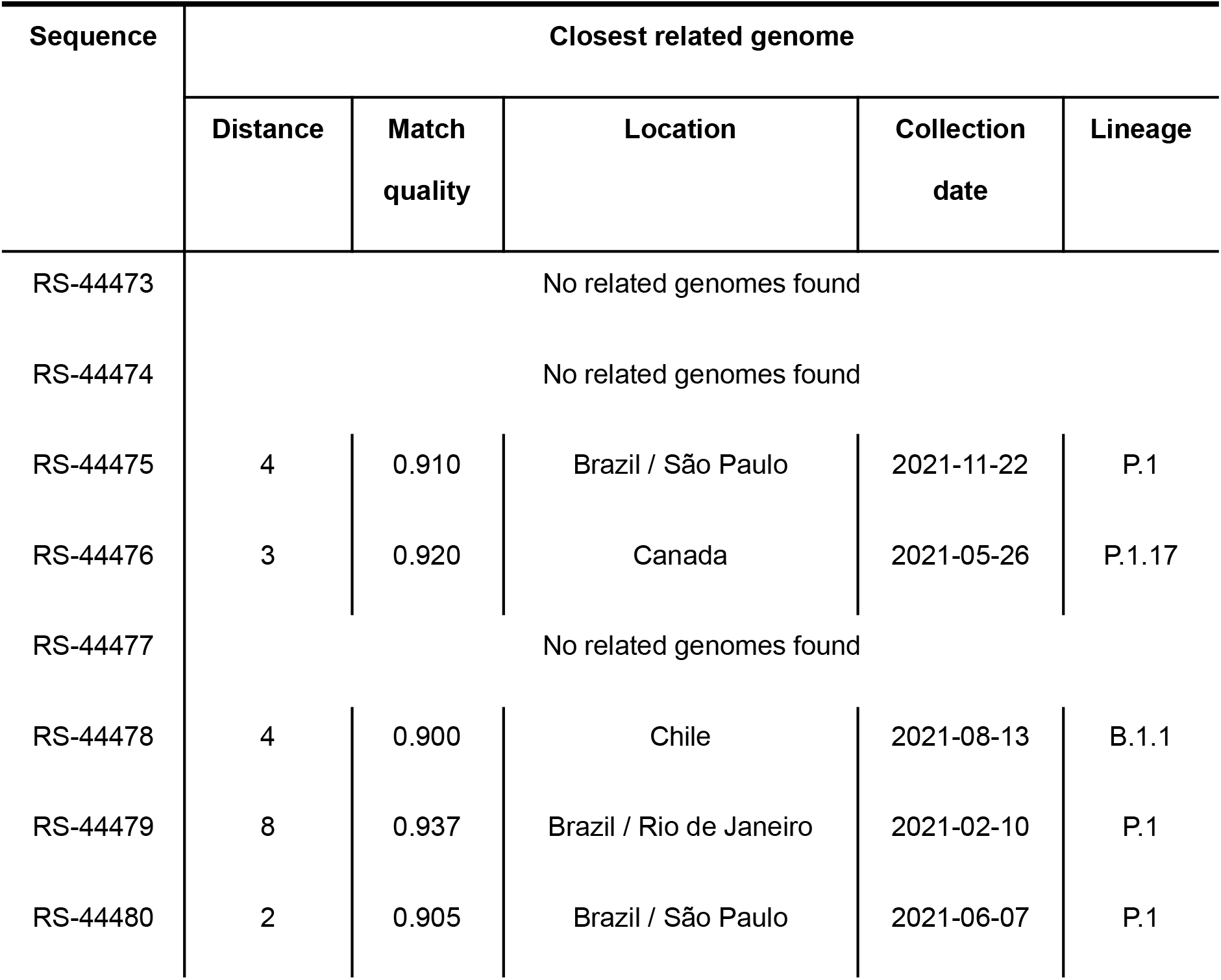
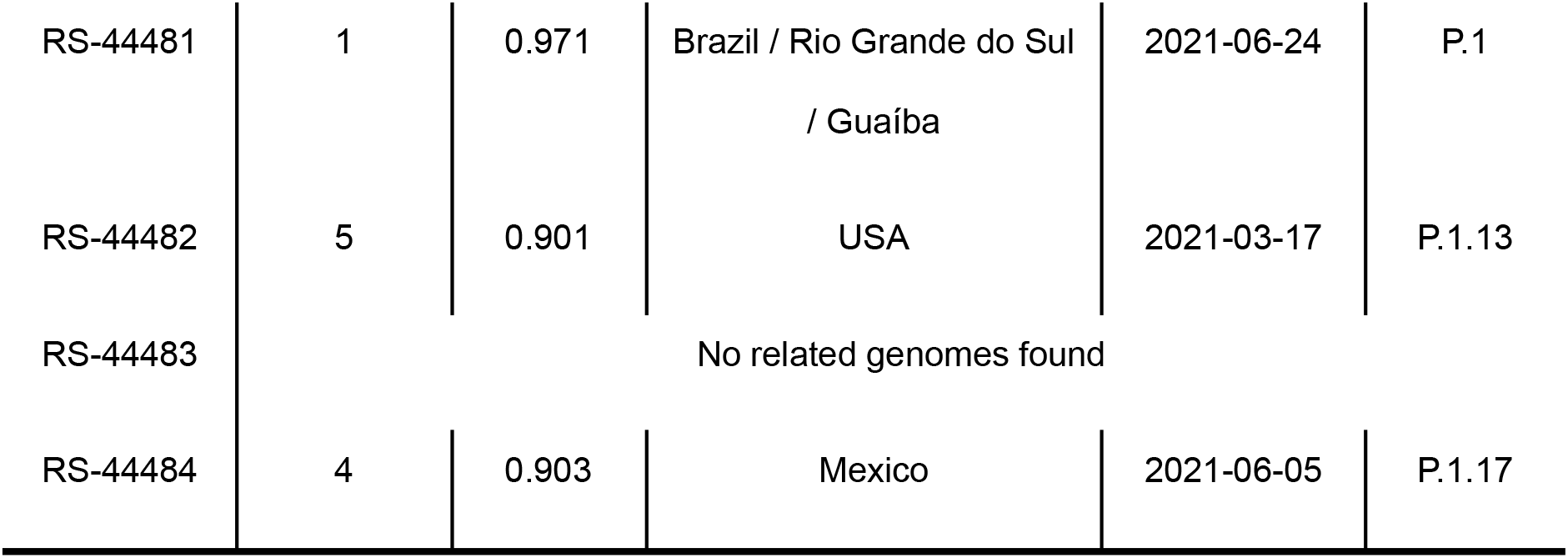
Closest related SARS-CoV-2 sequences from GISAID to the Esteio sequenced genomes from this study according to AudacityInstant search.

Besides the most closely related genomes, other 424 unique sequences (638 genomes in total) were recovered as being related to the sequenced genomes from this study with a genetic distance of 9 or less according to AudacityInstant parameters (Figure 4). Sequences RS-44475, RS-44478, RS-44479, and RS-44481 were mostly associated with Brazilian sequences (47.4 up to 89% of the retrieved genomes). RS-44476, RS-44480, and RS-44484 were predominantly related to Mexico (50%) and USA (30.3 and 27.5% of the genomes), respectively.

**FIGURE 4.**
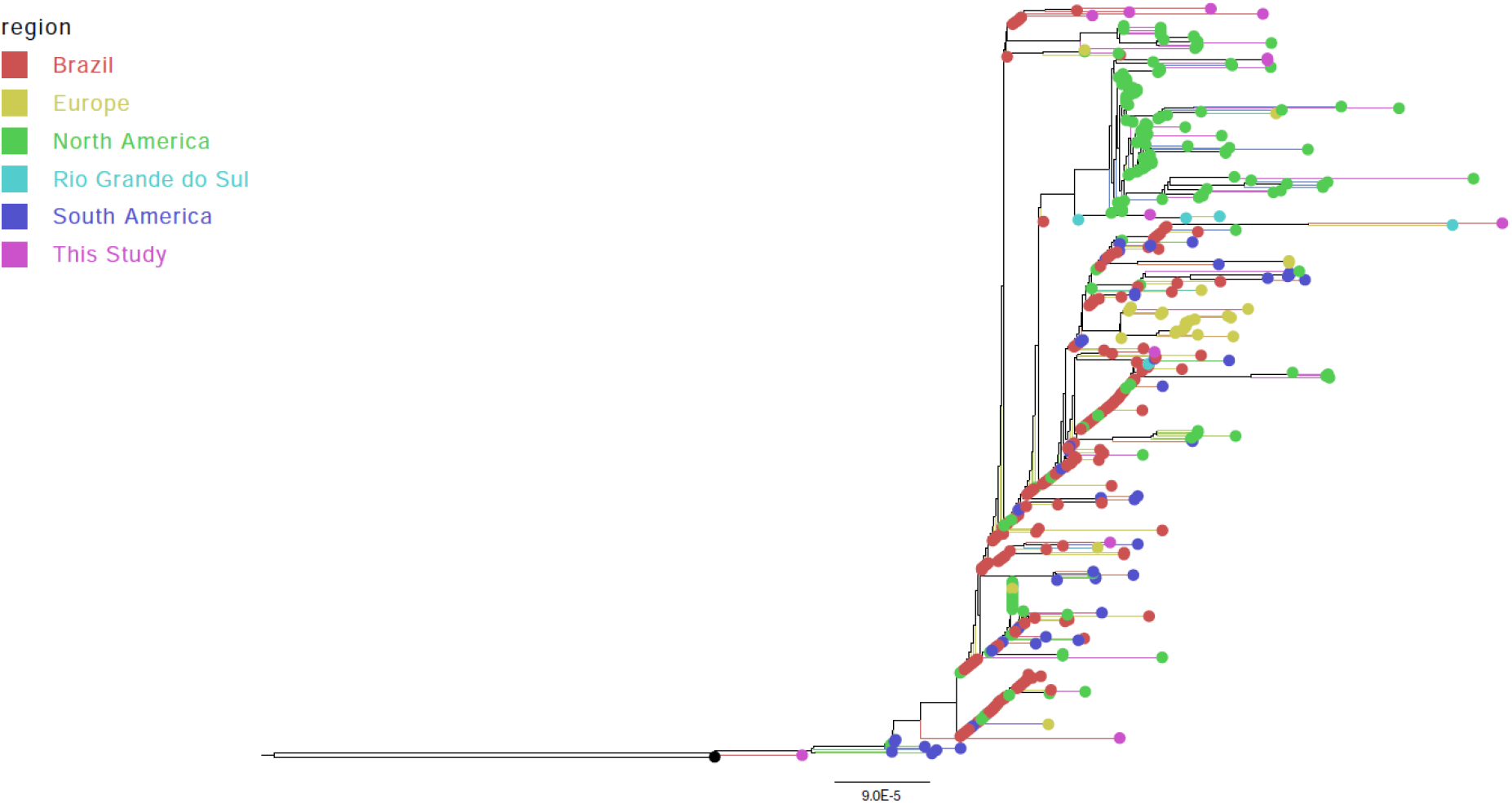
Global Maximum likelihood phylogenomic analysis with SARS-CoV-2 genomes closely related to eight sequenced genomes from this study according to Table 2.

### 2.5 Phylogenomic analysis of SARS-CoV-2 from Rio Grande do Sul state

The phylogenomic analysis of the SARS-CoV-2 genomes from Rio Grande do Sul state showed the formation of multiple monophyletic groups for the main VOCs (Figure 5). Alpha (B.1.1.7), Gamma (P.1 and derivative lineages), Delta (B.1.617.2 and derivative lineages), and Omicron (BA.2) clades were validated by SH-aLRT and aBayes tests with at least 97% of branch support (100/1, 99.9/1, 97.1/1, and 100/1 for Alpha, Gamma, Delta, and Omicron, respectively). For other lineages and former VOIs P.7 and Zeta (P.2), it was also observed the clustering in monophyletic groups (97.1/1, and 99/1 of statistical support for P.7 and P.2, respectively), as well as a larger clade including the P.1 (and its derivatives), P.2, and P.7 clades with B.1.1.28 sequences at basal branch (85.3/0.995 of statistical support for SH-aLRT and aBayes test). Interestingly, a clade with Alpha and Omicron genomes was formed with 86.2/0.996 of branch support.

**FIGURE 5.**
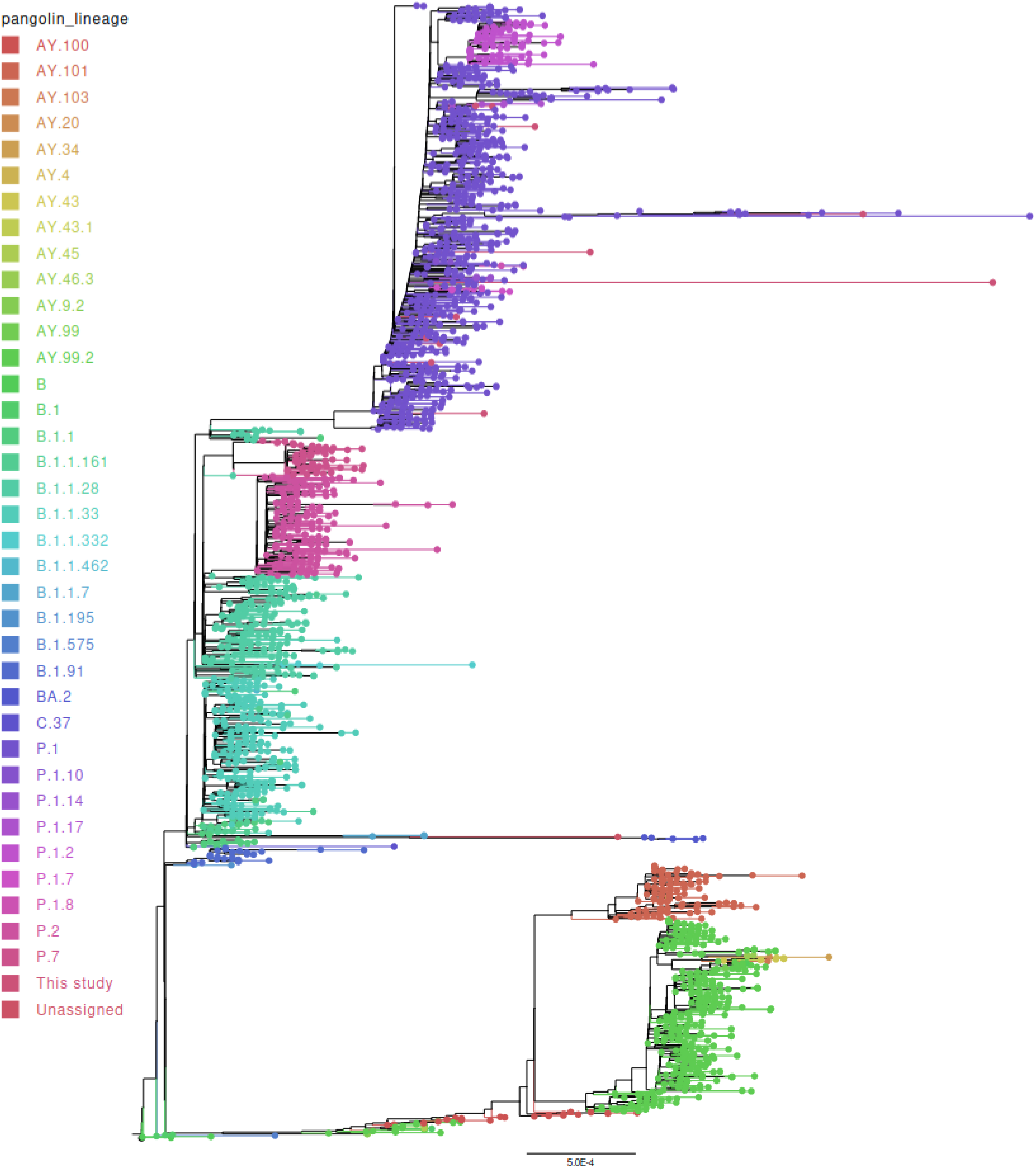
Maximum likelihood phylogenomic analysis of SARS-CoV-2 genomes from Rio Grande do Sul state.

As expected, all 12 genomes sequenced by this study clustered in the Gamma clade, which also presented subclades related to P.1 sublineages. P.1.2 (94/1 for SH-aLRT/aBayes), P.1.17 (85.3/0.997 for SH-aLRT/aBayes, including two genomes from this study at the basal branch), and P.1.7 (88.7/1 for SH-aLRT/aBayes) were found to form monophyletic groups. In the Delta group, sublineages AY.101 and AY.9.2 were supported as subclades by the statistical tests (96.3/1 and 100/1 for SH-aLRT/aBayes, respectively).

### 2.6 Phylogenetics and Molecular Evolution of SARS-CoV-2 Structural Proteins

The molecular evolutionary analysis aimed to identify positively and negatively selected sites from SARS-CoV-2 structural proteins in the genome dataset from Rio Grande do Sul state. The E, M, and N proteins were tested with HyPhy FUBAR, FEL, and SLAC methods (Tables 3 - 5).

**TABLE 3.** Protein E sites submitted to negative selection according to the HyPhy methods.

| Codon | FUBAR |  |  | FEL |  |  |  | SLAC |  |  |
| --- | --- | --- | --- | --- | --- | --- | --- | --- | --- | --- |
|  | Alpha | Beta | Post. prob. | Alpha | Beta | LRT | Prob. | dS | dN | Prob |
| 25 | -- | -- | -- | 9.477 | 0.000 | 2.747 | 0.0974 | -- | -- | -- |
| 63 | -- | -- | -- | 11.265 | 0.000 | 2.756 | 0.0969 | -- | -- | -- |

Post. prob.: Posterior probability. Prob: Probability. Gray rows indicate negative selection test results.

#### E protein

No sites were identified by FUBAR, FEL or SLAC methods to be under positive selective pressure in the Envelope protein. Two sites were identified by a negative selection test from the FEL method.

#### M protein

Two sites were identified to be under adaptive selection by FUBAR method in Membrane protein (Table 4). Twelve sites were found to be under purifying selection by FEL and/or SLAC methods, four of them by both methods.

**TABLE 4.** Protein M sites submitted to positive and negative selection according to the HyPhy methods.

| Codon | FUBAR |  |  | FEL |  |  |  | SLAC |  |  |
| --- | --- | --- | --- | --- | --- | --- | --- | --- | --- | --- |
|  | Alpha | Beta | Post. prob. | Alpha | Beta | LRT | Prob. | dS | dN | Prob |
| 53 | -- | -- | -- | 7.466 | 0.000 | 6.794 | 0.0091 | 3.447 | 0.000 | 0.024 |
| 94 | 0.764 | 6.559 | 0.9003 | -- | -- | -- | -- | -- | -- | -- |
| 112 | -- | -- | -- | 5.455 | 0.000 | 4.538 | 0.0332 | 2.306 | 0.000 | 0.084 |
| 114 | -- | -- | -- | 15.269 | 0.000 | 7.834 | 0.0051 | -- | -- | -- |
| 121 | -- | -- | -- | 5.333 | 0.000 | 2.799 | 0.0943 | 2.294 | 0.000 | 0.084 |
| 125 | 1.006 | 10.637 | 0.9555 | -- | -- | -- | -- | -- | -- | -- |
| 135 | -- | -- | -- | 11.754 | 0.000 | 3.455 | 0.0631 | -- | -- | -- |
| 138 | -- | -- | -- | 4.521 | 0.000 | 3.761 | 0.0524 | -- | -- | -- |
| 139 | -- | -- | -- | 7.642 | 0.000 | 3.612 | 0.0574 | -- | -- | -- |
| 147 | -- | -- | -- | 7.642 | 0.000 | 3.292 | 0.0696 | -- | -- | -- |
| 166 | -- | -- | -- | 12.036 | 0.000 | 3.279 | 0.0702 | -- | -- | -- |
| 195 | -- | -- | -- | 7.642 | 0.000 | 3.873 | 0.0491 | -- | -- | -- |
| 203 | -- | -- | -- | 8.065 | 0.000 | 4.217 | 0.0400 | 3.443 | 0.000 | 0.024 |
| 208 | -- | -- | -- | 7.764 | 0.000 | 3.503 | 0.0613 | -- | -- | -- |

Post. prob.: Posterior probability. Prob: Probability. Gray rows indicate negative selection test results.

#### N protein

Fourteen sites were identified to be under positive selective pressure by FUBAR and/or FEL in Nucleocapsid protein, of which nine were identified by both methods (Table 5). Forty-six sites were found to be under negative selective pressure, twenty of them identified by FEL and SLAC methods.

**TABLE 5.**
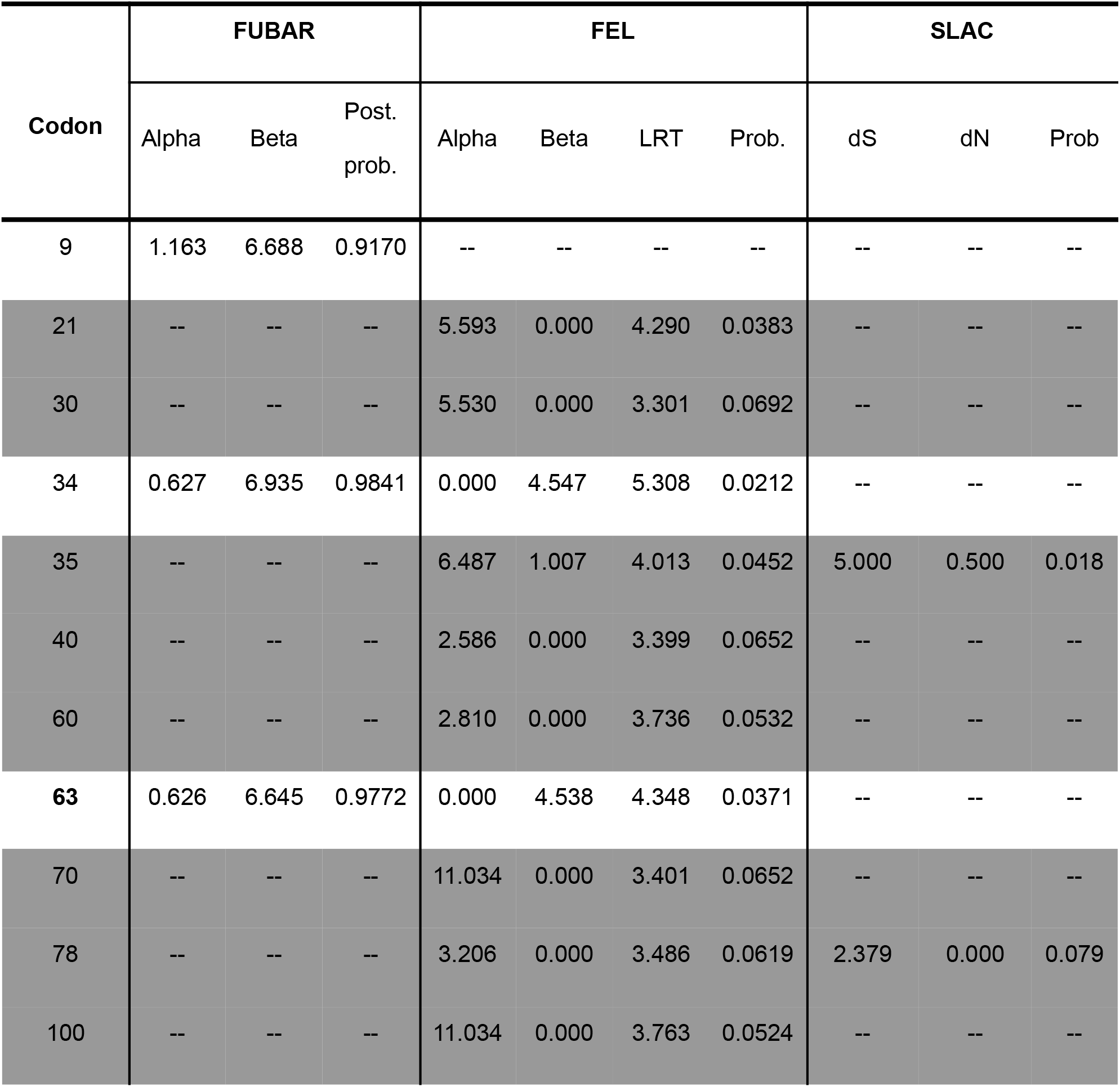
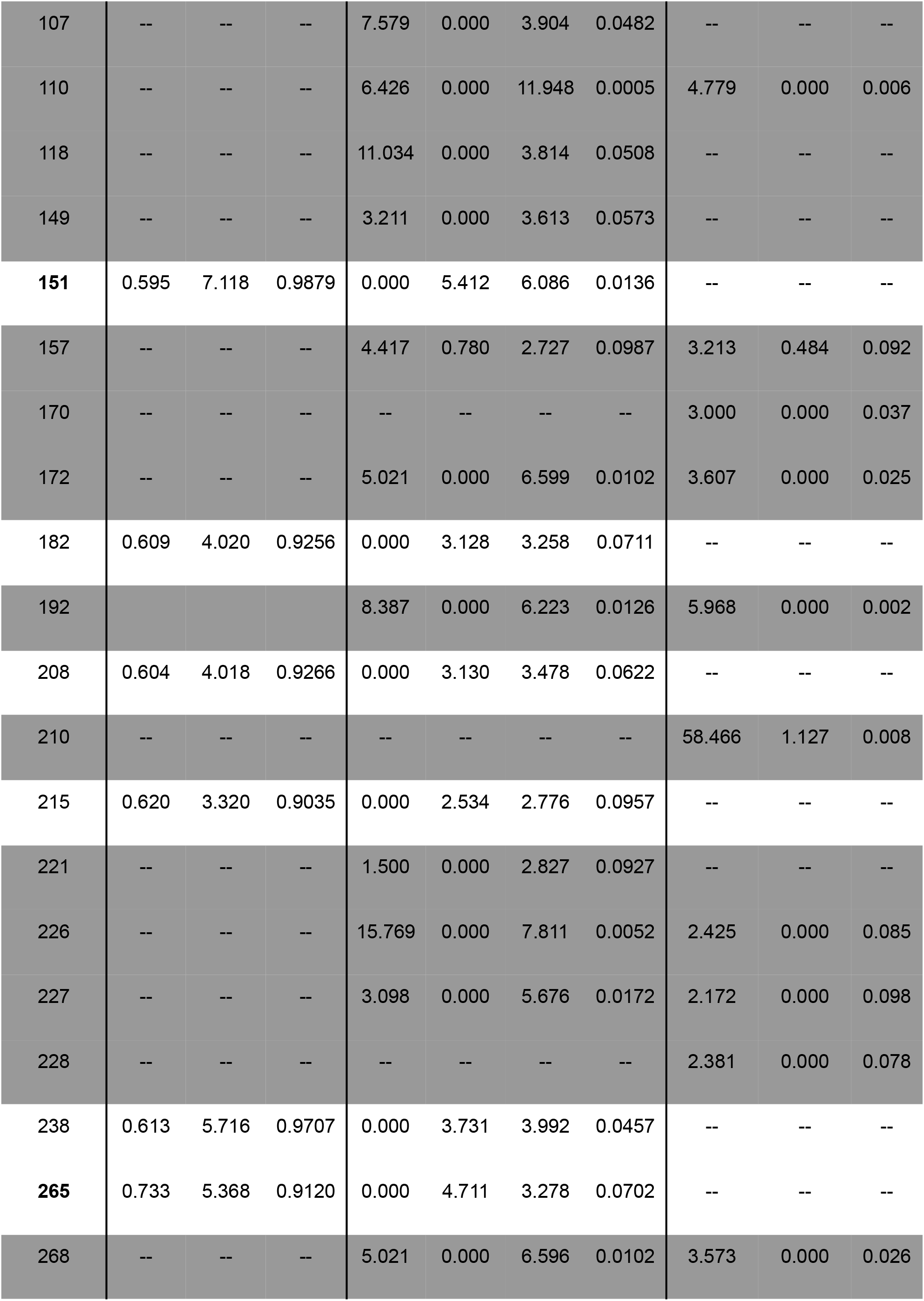
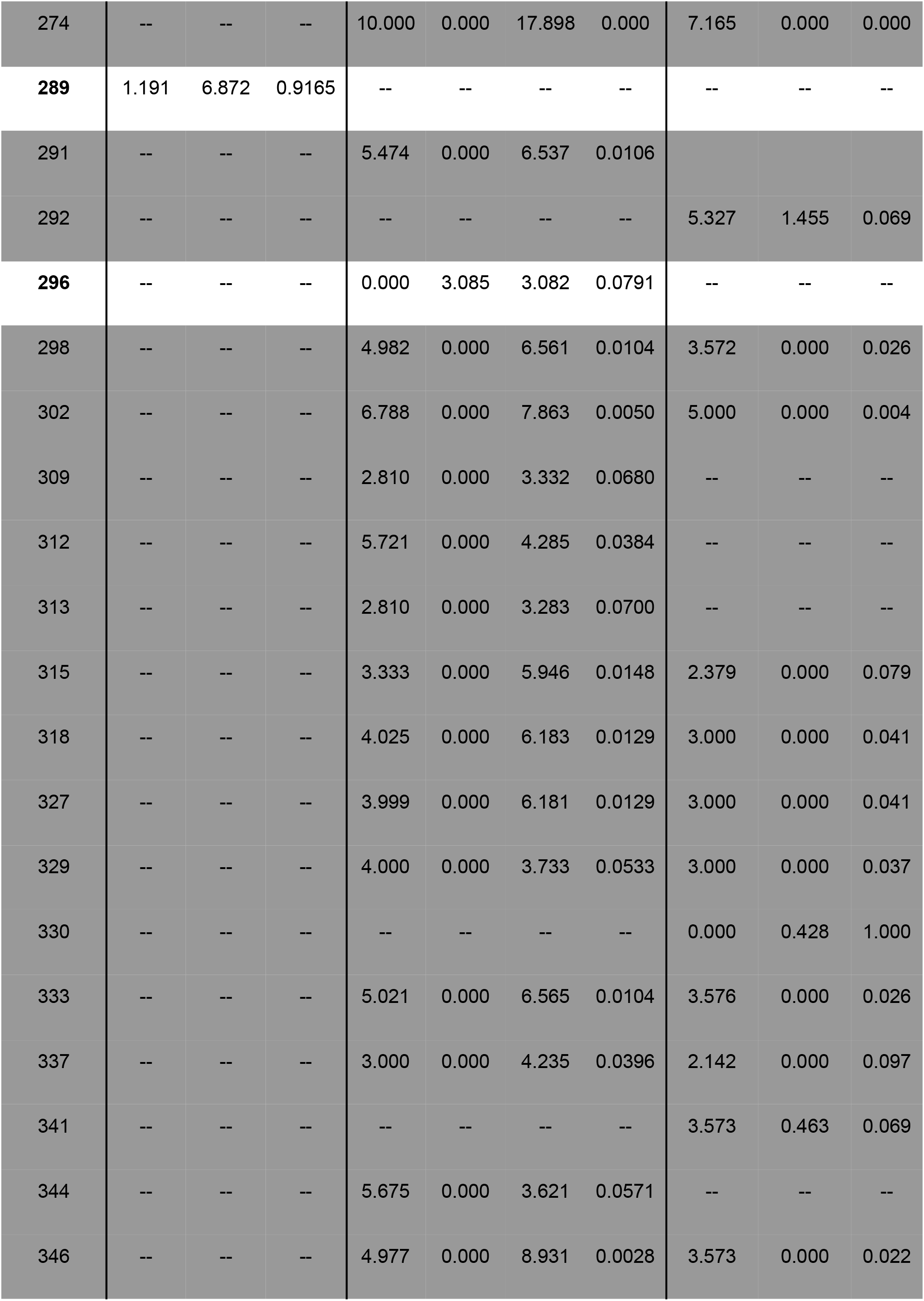
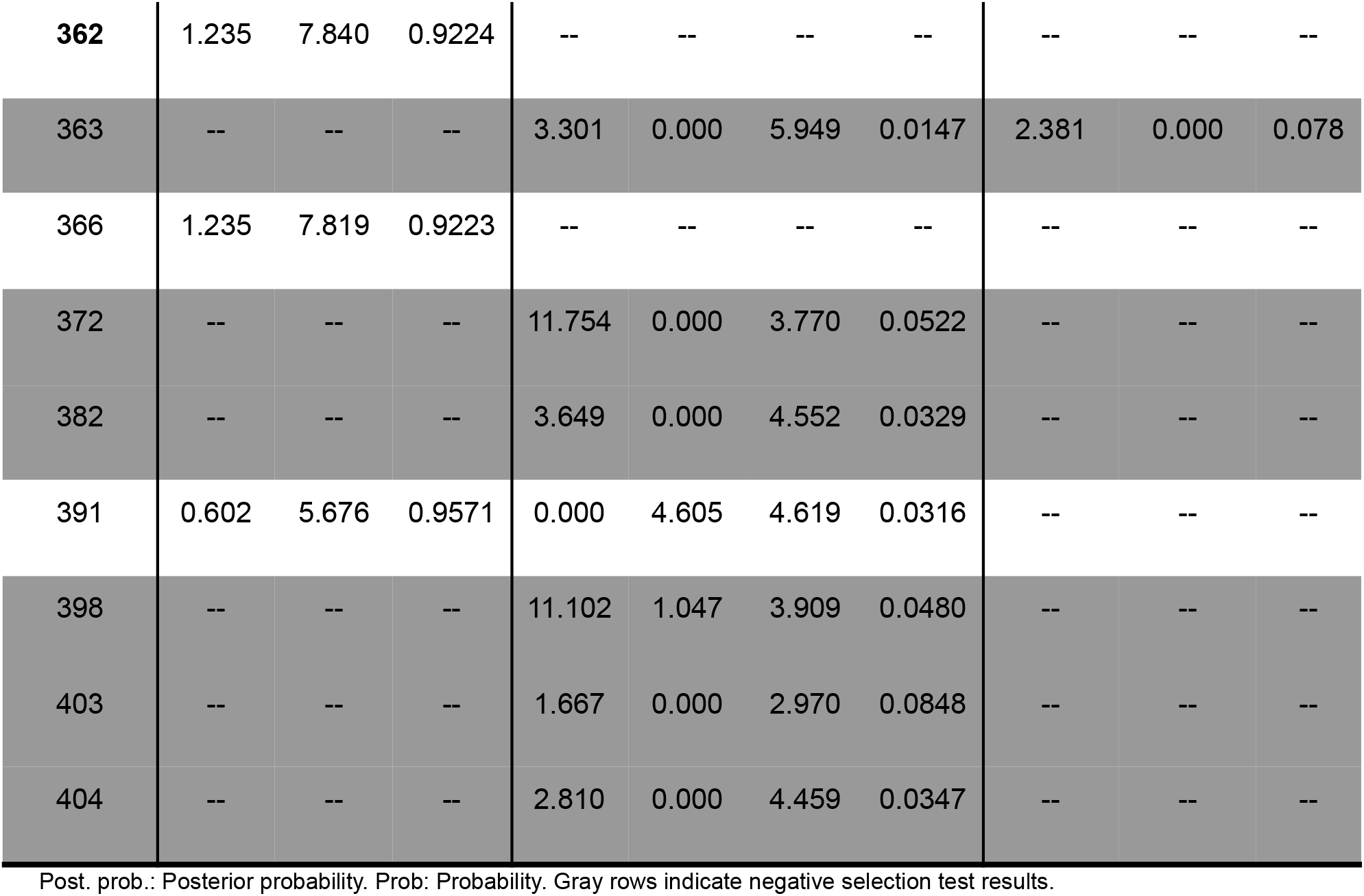
Protein N sites submitted to positive and negative selection according to HyPhy methods.

### 2.7 Molecular stability of the structural proteins E, M, and N

The program DynaMut2 was used to estimate the molecular stability of the SARS-CoV-2 structural proteins M and N with mutated residues at sites previously identified under positive selection (Table 6). Sites of M and N proteins recognized by the HyPhy tests had their amino acid substitutions evaluated at molecular level using publicly available structures from PDB database (Figure 6). Differently from the spike protein, proteins M and N are less represented in experimentally resolved structures from PDB. Thus, structures: (a) 8CTK - relative to protein M; (b) 7VNU - relative to N-terminal domain of protein N; and (c) 6ZCO - relative to C-terminal domain of protein N were selected to perform these analyzes.

**FIGURE 6.**
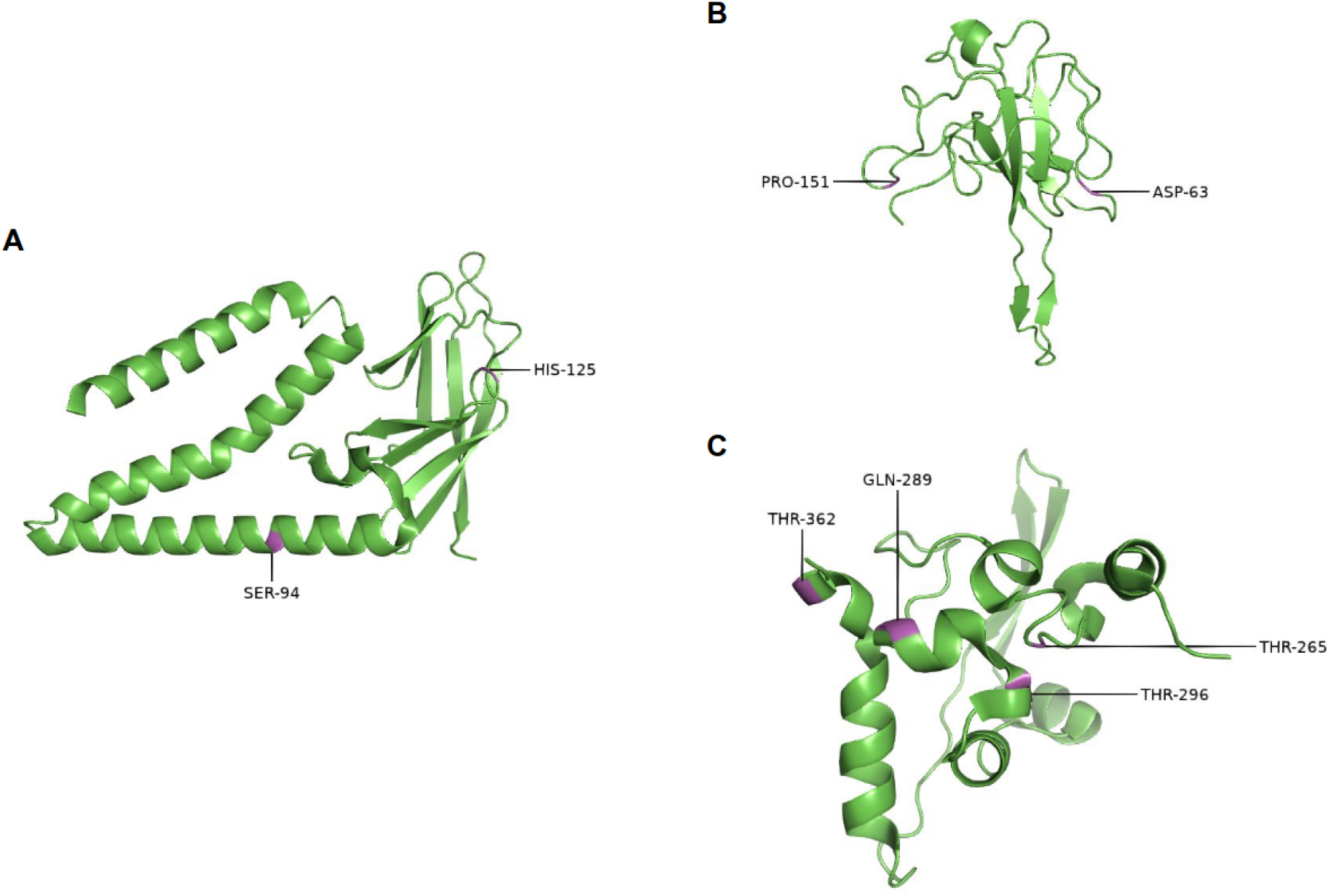
Positively selected sites in M and N protein structures. (A) Membrane protein (8CTK, chain A); (B) N-terminal domain from Nucleocapsid protein (7VNU, chain A); (C) C-terminal domain from Nucleocapsid protein (6ZCO, chain A).

**TABLE 6.** DynaMut2 results for positively selected sites from proteins E, M, and N.

| Protein | Site | Mutation | Frequency<sup>1</sup> | Predicted Stability Change ($\Delta\Delta G^{\text{Stability}}$) |
| --- | --- | --- | --- | --- |
| M | 94 | S → G | 0.45% | -0.19 kcal/mol (Destabilizing) |
| M | 94 | S → I | 0.09% | 0.65 kcal/mol (Stabilizing) |
| M | 125 | H → L | 0.04% | 1.83 kcal/mol (Stabilizing) |
| M | 125 | H → Q | 0.04% | 0.52 kcal/mol (Stabilizing) |
| M | 125 | H → Y | 0.18% | 2.04 kcal/mol (Stabilizing) |
| N | 63 | D → G | 18.53% | 0.08 kcal/mol (Stabilizing) |
| N | 63 | D → Y | 0.04% | 1.0 kcal/mol (Stabilizing) |
| N | 151 | P → L | 0.22% | -0.43 kcal/mol (Destabilizing) |
| N | 151 | P → S | 0.09% | -0.17 kcal/mol (Destabilizing) |
| N | 265 | T → I | 0.04% | -0.32 kcal/mol (Destabilizing) |
| N | 289 | Q → L | 0.04% | 0.42 kcal/mol (Stabilizing) |
| N | 289 | Q → H | 0.22% | -0.38 kcal/mol (Destabilizing) |
| N | 296 | T → I | 0.09% | -0.75 kcal/mol (Destabilizing) |
| N | 362 | T → I | 0.18% | -0.12 kcal/mol (Destabilizing) |
| N | 362 | T → K | 0.09% | -0.09 kcal/mol (Destabilizing) |

<sup>1</sup> Mutation frequency in the multiple sequence alignment (n = 2,240 sequences).

In the analysis of M protein, most alterations promote a stabilizing effect in the protein structure, excepting for S94G (ΔΔG = -0.19 kcal/mol). In the N-terminal domain from N protein, alterations in site 63 were associated with a stabilizing effect, while mutations in site 151 seems to destabilize the protein structure. Mutations observed in the C-terminal majoritarily suggest a destabilizing effect, excepting for mutation Q289L (ΔΔG = 0.42 kcal/mol). Some mutations such as M:S94G/I and N:Q289H/L showed variable stabilizing/destabilizing patterns for the same site. Mutations M:H125L/Y and N:D63Y not only presented a stabilizing effect but also are associated with a larger predicted stability change, increasing from 1 up to 2.04 kcal/mol in Gibbs Free Energy.

## 3 DISCUSSION

Rio Grande do Sul (RS) is currently the fourth state most affected by COVID-19 in Brazil (https://covid.saude.gov.br/ accessed on December 13, 2022). The P.1 lineage initiated a new wave of infections in Brazil around November 2020, starting in Manaus (northern Brazil) and spreading across the country (Faria et al., 2021). In RS, P.1 arrived in mid of January 2021, with a high transmission rate until April 2021, characterizing the second COVID-19 wave in the state (Varela et al., 2021; Salvato et al., 2021).

According to the phylogenomic analysis, the genomes from RS state formed monophyletic groups for most of the lineages, with specific clades to lineages and sublineages belonging to Alpha, Gamma, Delta, and Omicron VOCs. This data may suggest some intra-lineage genetic conservation in the SARS-CoV-2 genomes from RS state. However, as seen in AudacityInstant data search, four of our sequenced samples are most closely related to genomes from Chile, Mexico, Canada, and USA. It is not possible to guarantee if it is not an artifact from sequencing related to low quality and covering, sampling or if it is real evidence of possible migration events, since our genomes present older collection dates than their matches. The sequencing reads assembly for these four samples achieved between 98.12 and 99.73% of SARS-CoV-2 reference sequence covering and the occurrence of undefined nucleotides (Ns) ranged from 4.47 up to 17.78%, indicating low coverage sequences.

For those sequences more closely related to Brazilian samples, except for genome RS-44479 - which was collected in June, 2021 and had their closest related sequence dated to February, 2021, in Rio de Janeiro - the remaining ones also have older collection dates than their matches. The sequencing reads assembly for these four “Brazilian” samples achieved between 98.82 and 99.76% of SARS-CoV-2 reference sequence covering and the occurrence of undefined nucleotides (Ns) ranged from 2.00 up to 13.47%, indicating three low and one high coverage sequence in this set. The remaining four genomes with no identifiable related sequences presented very low sequencing quality and covering, ranging from 84.92 up to 98.26% of SARS-CoV-2 reference sequence covering and 49.55 up to 58.43% of undefined nucleotides.

RNA viruses have a higher mutation rate than DNA viruses and organisms (Duffy, 2018). Selective pressure occurs in a way that the virus can keep its transmission and immune evasion mechanisms updated according to the host characteristics (Zarai et al., 2020). The E protein is the smallest structural protein of SARS-CoV-2 and keeps their structure highly conserved across diverse genres of β-coronaviruses (Yadav et al., 2021). It comprises three main domains, the N-terminal (NT), C-terminal (CT), and transmembrane domain (TMD). Possible modification in TMD could indicate an differential interaction with membrane lipids, as well as the alteration of the capacity of membrane attachment and ER targeting by the E protein (Timmers et al., 2021). Similarly, sites located at the D-L-L-V motif bind to the host protein PALS1 could facilitate infection (Timmers et al., 2021). However, no sites were found under positive selective pressure in E protein. The presence of sites 25 and 63 under negative selective pressure suggest their importance to protein function conservation.

The M protein is very important in the mounting of the virion and the other structural proteins in the coronaviruses (Neuman et al., 2011). In SARS-CoV-2, this protein can be related to antigenic reactions, with the S and N proteins (Lopandić et al., 2021) even reducing the interferon I responses (Sui et al., 2021). Therefore, modifications in its genomic structure can directly impact the virus survival, which is probably the reason for the low identification of diversifying selection events in that protein.

The N protein structure is composed of three main domains: N-terminal domain (NTD), a linker domain rich on serine and arginine residues (SR-rich linker), and a C-terminal domain (CTD) (Timmers et al., 2021). NTD and CTD comprise major antigenic sites of the N protein in SARS-CoV virus (Surjit & Lal, 2009). This protein has a role in the packing of the viral genetic material besides related to immune escape, blocking interferons and other defense mechanisms of the host (Bai et al., 2021). According to Rahman et al., several alterations in that protein makes it difficult to create vaccines and medications that could use it as a target (Rahman et al., 2020). The co-occurring amino acid mutations R203K and G204R, for example, are known to enhance replication, fitness, and pathogenesis of SARS-CoV-2 (Johnson et al., 2022).

Changes in nucleotides can result in modification of the protein structure, increasing or decreasing their stability (Jaenicke, 1996). The flexibility of a protein is related to its function and conformation (Zhao, 2010). In this way, the supervised machine-learning trained tool DynaMut2 was selected to predict the effect of missense variations from positively selected sites on protein stability. According to the DynaMut2 results for M protein, mutations in site 94 could stabilize or destabilize protein structure according to the alteration of the native serine by an isoleucine or a glycine, respectively. Site 125 achieved a stabilizing effect in all tested mutations. The mutation H125Y is the most frequent on GISAID with 14,355 occurrences in SARS-CoV-2 genomes in the world (accessed on October 31, 2022). Present in all variants of concern, this mutation was found to be prevalently associated to the Delta and Omicron clades, 35.10% and 24.46% of the occurrences on GISAID, respectively, while the variant H125L is less spread, occurring in ≅0.001% of world genomes, including those from Alpha, Delta, and Omicron groups. Another minor variant, H125Q (≅0.0009%) is also related to Alfa, Delta, and Omicron lineages.

The N-terminal domain of N protein had two sites analyzed with 2 different mutations each. Mutations on site 63 lead to a stabilizing effect and mutations on site 151 seem to destabilize protein structure. The alteration of an aspartic acid to a glycine in D63G mutation is widely found in the world, occurring in 32.74% of world genomes on GISAID, majoritarily in Delta sequences (99.8% of D63G occurrences). D63Y is found in approximately 2,800 genomes, mostly from Omicron and Alpha lineages. P151S substitution (≅1.28% of SARS-CoV-2 genomes in the world) destabilizes N-terminal domain from Nucleocapsid protein by alteration of a non-polar proline by a polar serine. Prevalent in Omicron (95.64% of P151S occurrences on GISAID), P151S is more frequent than P151L (non-polar proline to non-polar leucine), which destabilizes the protein structure, with more than 26,000 occurrences on SARS-CoV-2 genomes in the world, including all VOCs.

In the C-terminal domain of N protein were considered four sites with their respective amino acid alterations and the majority of them seem to destabilize the protein. Only mutation Q289L (≅0.009% of GISAID genomes) demonstrates a stabilizing effect on the C-terminal domain structure. Surprisingly, as observed in N-terminal domain mutations, Q289H (≅0.08% of GISAID genomes in the world) that lead to a destabilizing effect is nine times more frequent than the substitution for a leucine residue and is mostly found in Delta and Omicron genomes.

Similar results were found by Rahman and colleagues (2020) in the analysis of the structural effects of mutation Q289H. For T362I, their results indicate a stabilizing effect, in contrast with our findings. Finally, more studies are necessary to completely understand how structural changes may lead to advantages of SARS-CoV-2 in the host-pathogen interactions.

## 4 MATERIALS AND METHODS

### 4.1 Sample collection and clinical testing

Respiratory secretion were analyzed by Laboratório Central de Saúde Pública do Estado do Rio Grande do Sul (LACEN) (Porto Alegre, Rio Grande do Sul, Brazil) using RT-qPCR AllPlex SARS-CoV-2 assays Seegene Inc. Seoul, Republic of Korea with primers and probes targeting the RNA dependent RNA Polymerase (RdRP) Nucleocapsid (N) and Envelope (E) genes as recommended by the World Health Organization, with remnant samples stored at -20ºC. For the sequencing protocol, positive samples in the first RT-qPCR between April 09, 2021 to June 29, 2021, were selected and submitted to a second RT-qPCR, which was performed by BiomeHub (Florianópolis, Santa Catarina, Brazil), with a charite-berlin protocol. Samples with quantification cycle (Cq) up to 30 for at least one primer were selected for SARS-CoV-2 genome sequencing and assembly by the BiomeHub laboratory. In total, 12 samples who tested positive for SARS-CoV-2 RT-qPCR were included in the study.

### 4.2 SARS-CoV-2 genome sequencing and assembly

Total RNAs were prepared according to a reference protocol (Eden & Sim, 2020), with cDNA synthesized with SuperScript IV (Invitrogen) and DNA amplified with Platinum Taq High Fidelity (Invitrogen). The library preparation was performed with Nextera Flex (Illumina) and quantification was performed with Picogreen and Collibri Library Quantification Kit (Invitrogen). The genome sequencing was generated on Illumina MiSeq Platform by 150×150 runs with 500xSARS-CoV-2 coverage (50-100 mil reads/per sample).

For the genome assembly (BiomeHub in-house script), the adapters removal and read trimming for 150 nt read sequences were performed by fastqtools.py. The alignment of the sequenced reads to the reference SARS-CoV-2 genome (GenBank ID: NC_045512.2) was performed by Bowtie v2.4.2 (Langmead & Salzberg, 2012) with additional parameters as end-to-end and very-sensitive. The analyses of the sequencing coverage and depth were generated by samtools v1.11 (Li et al., 2009) with minimum base quality per base (Q) ≥ 30. Finally, the consensus sequence for each SARS-CoV-2 genome was generated by a bcftools pipeline (Li, 2011), including the commands mpileup (parameters: Q ≥ 30; q ≥ 40, depth (d) ≤ 2,500), filter (parameters: DP>50), call and consensus.

### 4.3 SARS-CoV-2 genomes and data retrieval

In order to compare 12 SARS-CoV-2 genomes from Esteio to other samples from the state, we gathered 2,227 sequences from the GISAID database (Elbe & Buckland-Merrett, 2017) with a collection date between March 1, 2020 and May 27, 2022 (submission up to May 27, 2022). The sequences were selected according to the following filters: (i) Location: South America / Brazil / Rio Grande do Sul; (ii) Clade: all; (iii) Complete genomes; (iv) High Coverage Selected.

The analysis of sequencing efforts, lineage frequencies and genomic characterization in Rio Grande do Sul state was posteriorly performed with GISAID data by retrieval of 4,706 sequences included in the following parameters: (i) Location: South America / Brazil / Rio Grande do Sul, (ii) Collection date between March 1, 2020 and May 31, 2022, and (iii) Submission date up to September 30, 2022.

### 4.4 SARS-CoV-2 mutations and lineages

SNPs and insertions/deletions in each sample were identified by the variant calling pipeline (https://github.com/tseemann/snippy), which uses FreeBayes and snpEff to call, annotate and predict variant effects on genes and proteins. The genomes were aligned with MAFFT and the extraction of SNPs and gaps from the sequences in relation to the reference was performed with msastats.py script. The reference sequence comes from the GenBank RefSeq (NC_045512.2), isolated and sequenced from an initial case from Wuhan, China, in 2019. The strains were identified using the dynamic nomenclature implemented in Pangolin (Rambaut, 2020) (https://github.com/cov-lineages/pangolin) and global clades and mutations using Nextstrain from Nextclade (https://clades.nextstrain.org/).

### 4.5 Phylogenomic analyses

For the global phylogenomics, a search was performed by Audacity *Instant* on the GISAID database (Elbe & Buckland-Merrett, 2017) to find closely related sequences to the sequenced genomes from this study (up to June 23, 2022). The resulting genome set was aligned with the MAFFT v.7 web server (Katoh et al., 2017). The trimming of 5’ and 3’ UTRs was performed with UGENE (Okonechnikov et al., 2012). The evolutionary model and phylogenomic tree inferences were performed by the IQ-TREE software (Nguyen et al., 2014) with addition of a Shimodaira-Hasegawa-like approximate likelihood ratio test of 1,000 replicates (Guindon et al., 2010) and an approximate Bayes test (Anisimova et al., 2011). Figtree software (http://tree.bio.ed.ac.uk/software/figtree/) was used to inspect and visualize the phylogenomic tree.

For the local phylogenomic analyses, 2,227 genome sequences from Rio Grande do Sul state, previously downloaded from the GISAID database, were aligned using the MAFFT v.7 web server. The trimming of 5’ and 3’ UTRs was performed with UGENE, identification of the best evolutionary model and phylogenomic inference performed by IQ-TREE software with the same parameters used for the global phylogenomics previously described, with Figtree software used for the inspection and visualization of phylogenomic tree.

### 4.6 Phylogenetics and Molecular Evolution of SARS-CoV-2 Structural Proteins

In order to infer the phylogenetic patterns of structural proteins E, M and N, genomic alignment coordinates related to these sequences were exported according to SARS-CoV-2 reference genome (NC_045512.2). Sequences with nucleotide insertions altering the reading frame were excluded from the analysis. For each gene sequence alignment, the evolutionary model and phylogenetic tree were inferred according to the previously described parameters from phylogenomic analysis.

Molecular evolution tests were performed with the HyPhy package (Pond et al., 2004). The methods FUBAR (Murrell et al., 2013), FEL (Kosakovsky Pond & Frost, 2005), and SLAC (Kosakovsky Pond & Frost, 2005) were implemented to evaluate potential sites under adaptive (pervasive) and purifying selection.

### 4.7 Molecular stability of structural proteins M and N

The estimation of molecular stability of structural proteins was performed by DynaMut2 web server (Rodrigues et al., 2020) using the experimentally resolved structures by Electron Microscopy: (a) 8CTK (3.52 Å) - relative to protein M; and X-Ray Diffraction: (b) 7VNU (1.95 Å) - relative to N-terminal domain of protein N; and (c) 6ZCO (1.36 Å) - relative to C-terminal domain of protein N, from Protein Data Bank (https://www.rcsb.org/). The selection of tested amino acid mutations was defined according to the sites detected under positive selection by the molecular evolution tests.

## Data Availability

Full tables acknowledging the authors and corresponding labs submitting sequencing data used in this study can be found in Supplementary File 2. The genomes sequenced by this study are deposited on GISAID database under identification codes EPI_ISL_16106069 up to EPI_ISL_16106069. Additional information related to the current study as well as the two genome sequences with long Ns stretches (>50%) are available from the corresponding author on reasonable request.

## CONFLICT OF INTEREST

The authors declare no conflict of interests.

## ETHICS STATEMENT

The research protocol was approved with exemption of written informed consent for viral genome sequencing and bioinformatic analyses by Comitê de Ética em Pesquisa em Seres Humanos of Universidade Federal de Ciências da Saúde de Porto Alegre (CEP - UFCSPA) under process number CAAE 39247920.0.0000.5345.

## AUTHOR CONTRIBUTION

**Amanda de M. Mayer:** Formal analysis; investigation; methodology; writing – original draft; writing - review and editing. **Patrícia A. G. Ferrareze:** Conceptualization, formal analysis; investigation; methodology; writing – original draft; writing - review and editing. **Luiz F. V. de Oliveira:** Methodology; resources; funding acquisition; writing - review and editing. **Tatiana S. Gregianini:** Methodology; resources; writing - review and editing. **Carla L. A. M. N**.: Resources; writing - review and editing. **Gabriel D. Caldana:** Investigation; writing – original draft; writing - review and editing. **Lívia Kmetzsch:** Supervision; writing - review and editing. **Claudia E. Thompson:** Conceptualization, formal analysis; investigation; methodology; resources; supervision; funding acquisition; writing – original draft; writing - review and editing. All authors have read and approved the manuscript.

## ACKNOWLEDGEMENTS

We thank the administrators of the GISAID database and research groups across the world for supporting the rapid and transparent sharing of genomic data during the COVID-19 pandemic and the *Governo do Estado do Rio Grande do Sul* and *Ministério da Saúde* for supplies and equipment used in the SARS-CoV-2 diagnosis routine. This study was financed in part by the *Coordenação de Aperfeiçoamento de Pessoal de Nível Superior - Brasil (CAPES)* - Finance code 001. The genome sequencing was performed by BiomeHub laboratory.

## Notes

### Competing Interest Statement

The authors have declared no competing interest.

### Funding Statement

This study was financed in part by the Coordenacao de Aperfeicoamento de Pessoal de Nivel Superior - Brasil (CAPES) - Finance code 001.

### Author Declarations

Comite de Etica em Pesquisa em Seres Humanos of Universidade Federal de Ciencias da Saude de Porto Alegre (CEP - UFCSPA) gave ethical approval for this work

