## Supplementary File 1 for "Genomic characterization and molecular evolution of SARS-CoV-2 in Rio Grande do Sul State, Brazil"

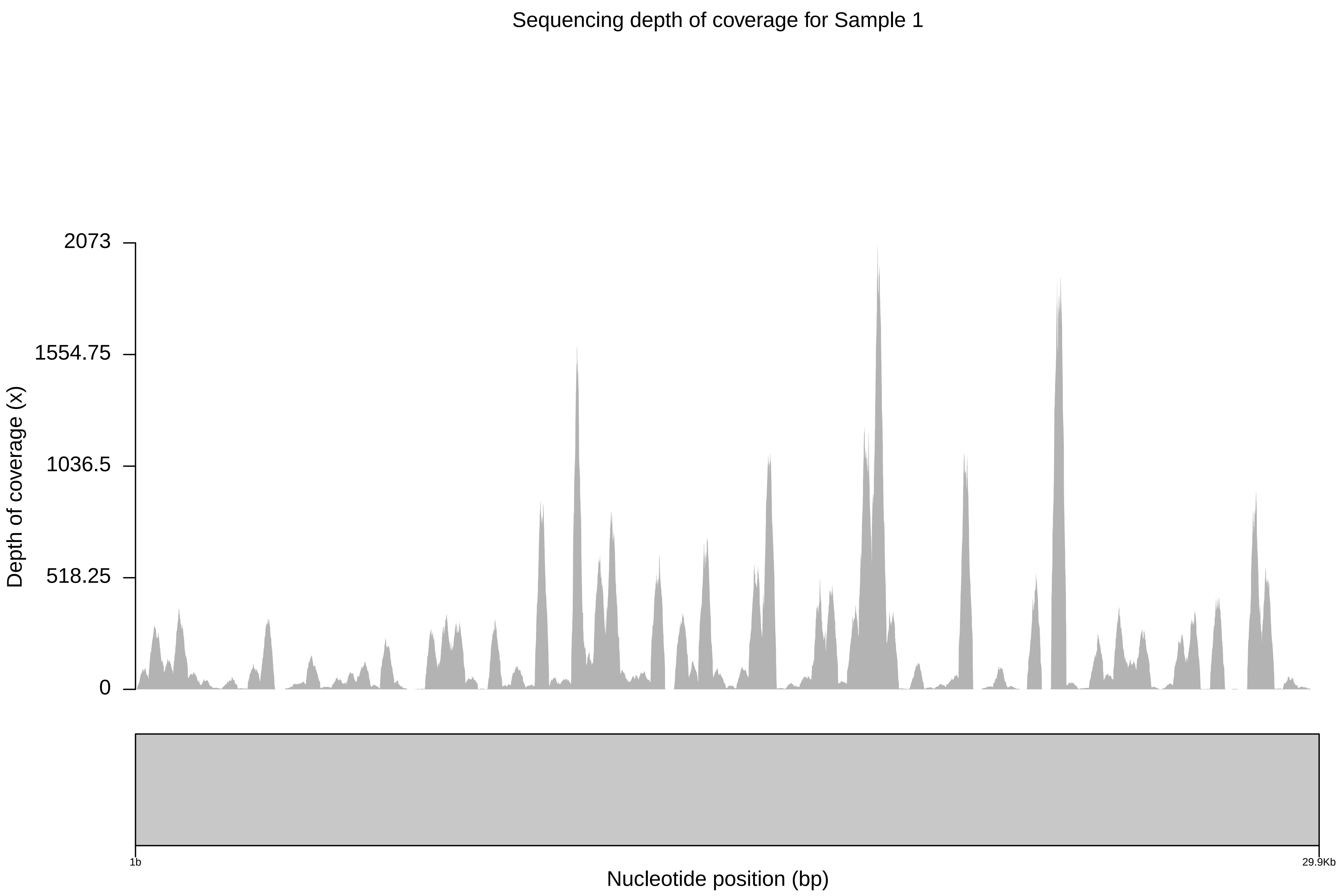
Sequencing depth of coverage for Sample 1
Depth of coverage (x)
2073
1554.75
1036.5
518.25
0
1b
29.9Kb
Nucleotide position (bp)

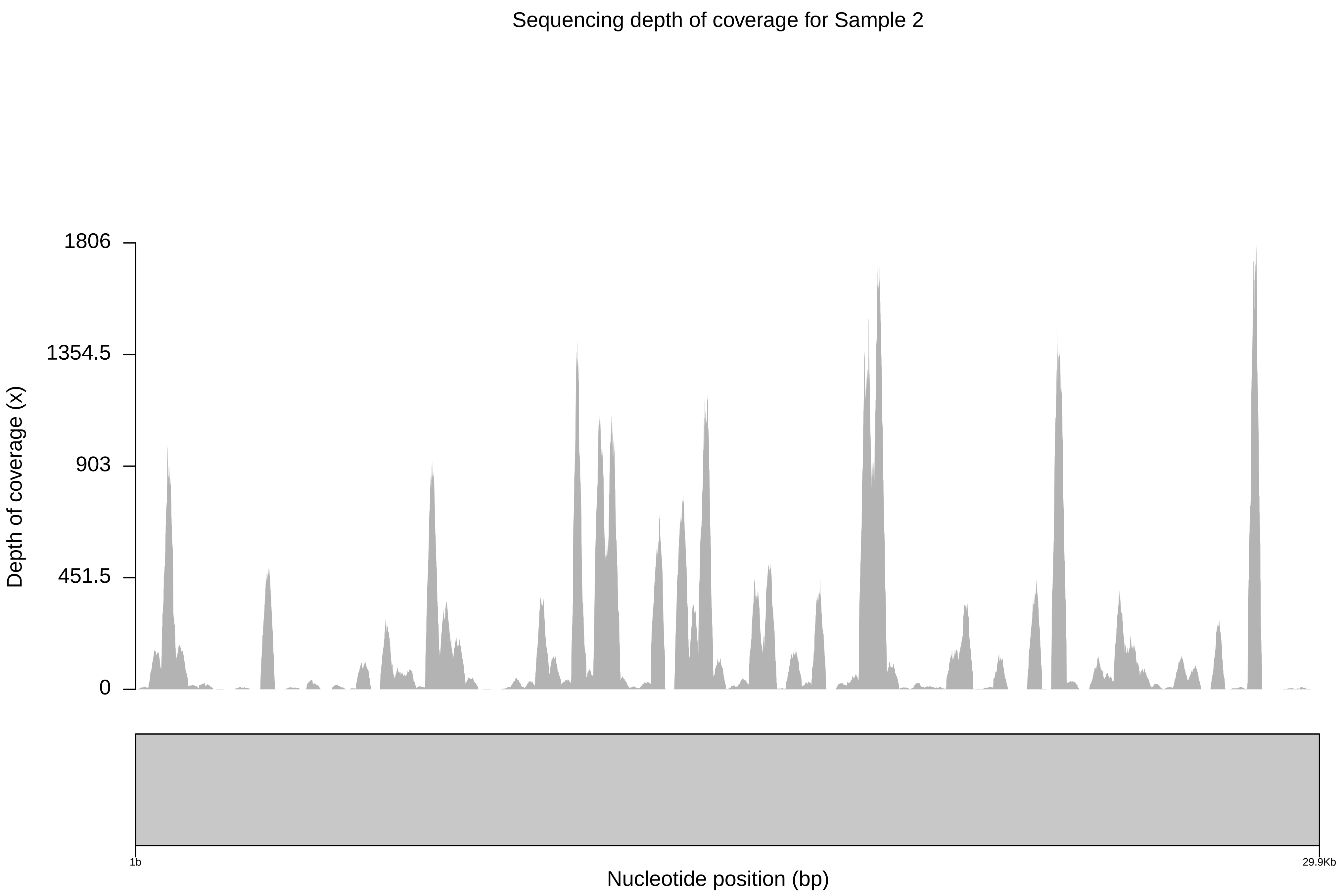
Sequencing depth of coverage for Sample 2
1806
1354.5
903
451.5
0
Depth of coverage (x)
1b
29.9Kb
Nucleotide position (bp)

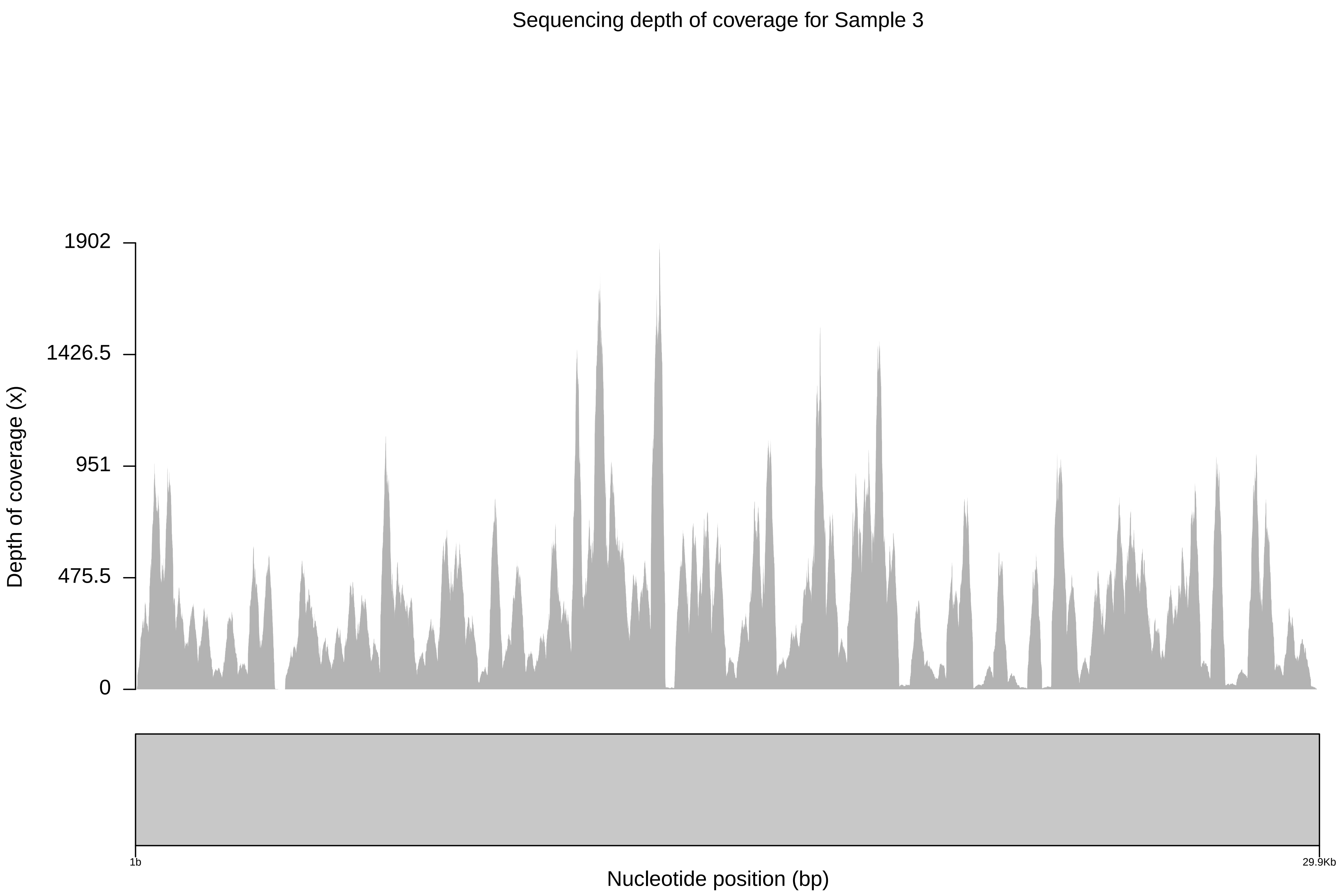

Sequencing depth of coverage for Sample 3
1902
1426.5
951
475.5
0
Depth of coverage (x)
1b
29.9Kb
Nucleotide position (bp)

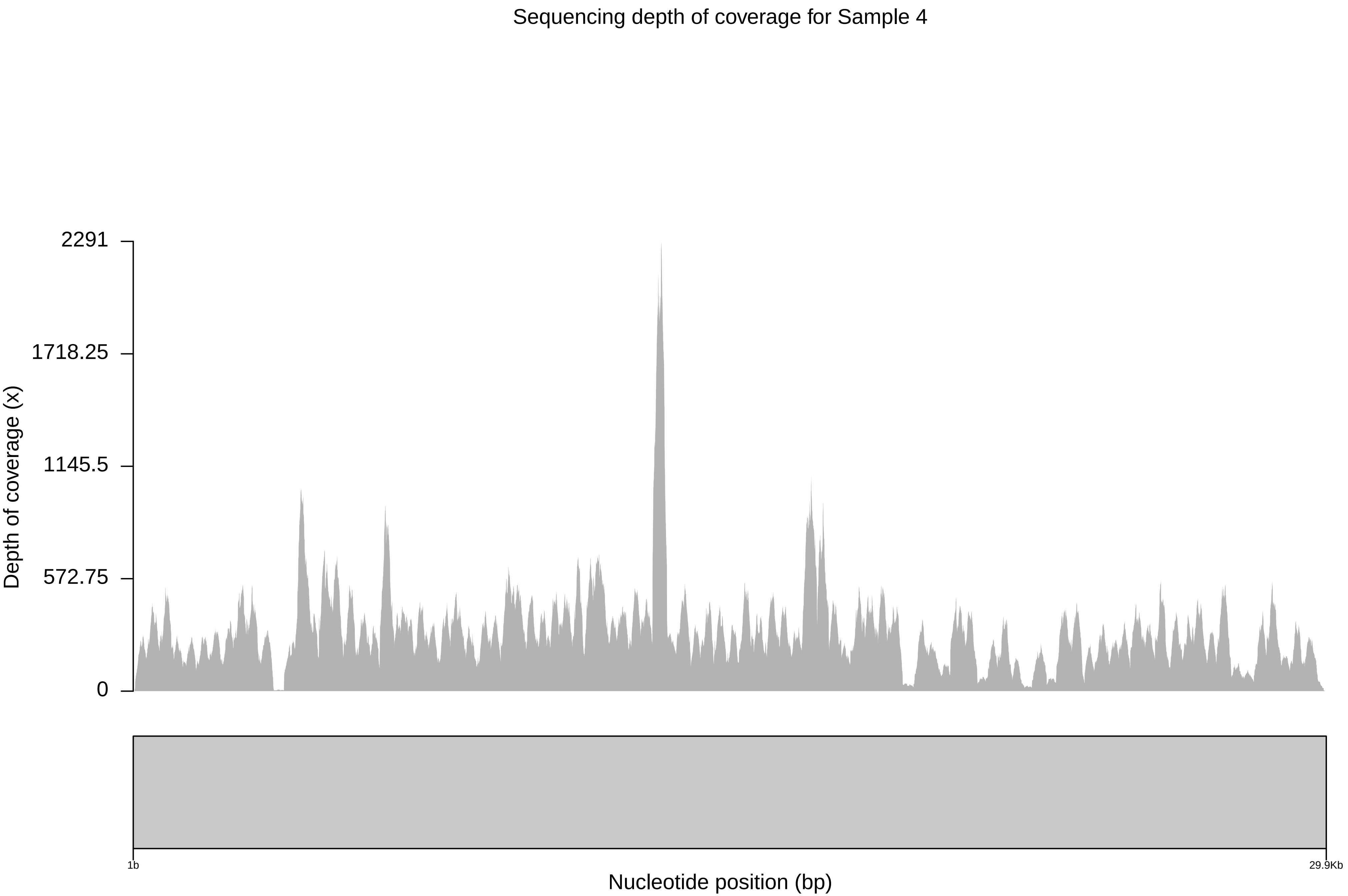

Sequencing depth of coverage for Sample 4
2291
1718.25
1145.5
572.75
0
Depth of coverage (x)
1b
29.9Kb
Nucleotide position (bp)

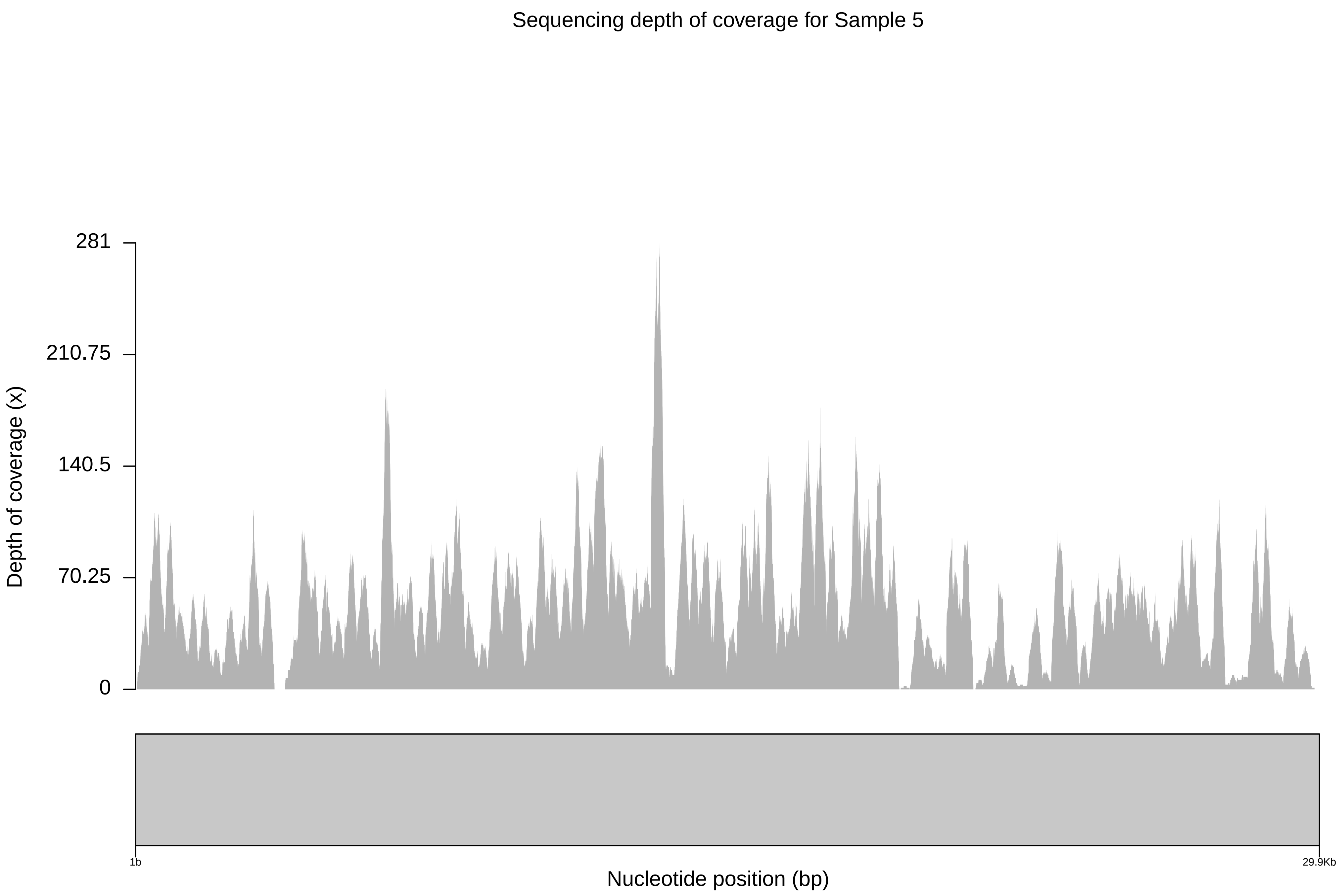

Sequencing depth of coverage for Sample 5
Depth of coverage (x)
281
210.75
140.5
70.25
0
1b
29.9Kb
Nucleotide position (bp)

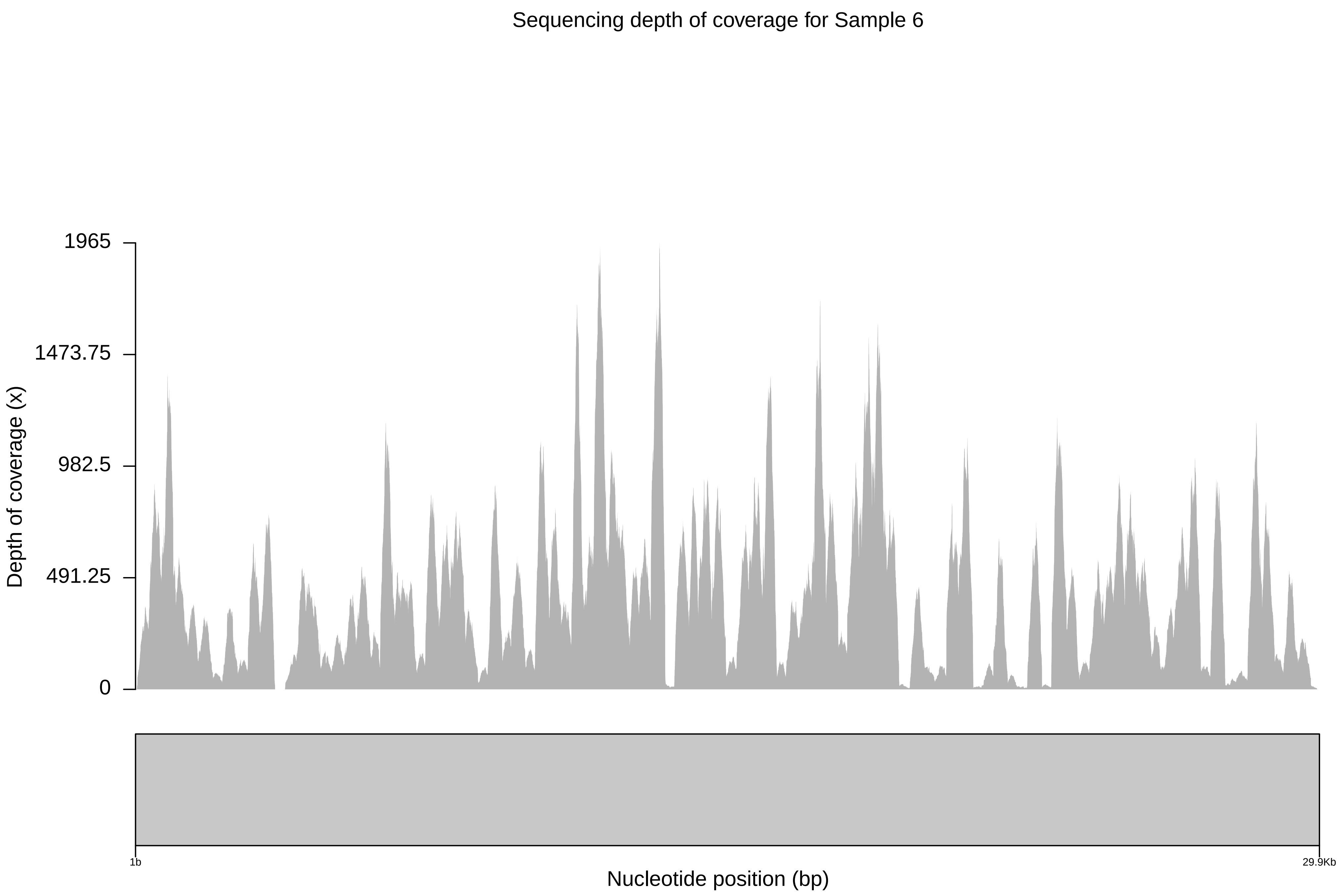

Sequencing depth of coverage for Sample 6
1965
1473.75
982.5
491.25
0
Depth of coverage (x)
1b
29.9Kb
Nucleotide position (bp)

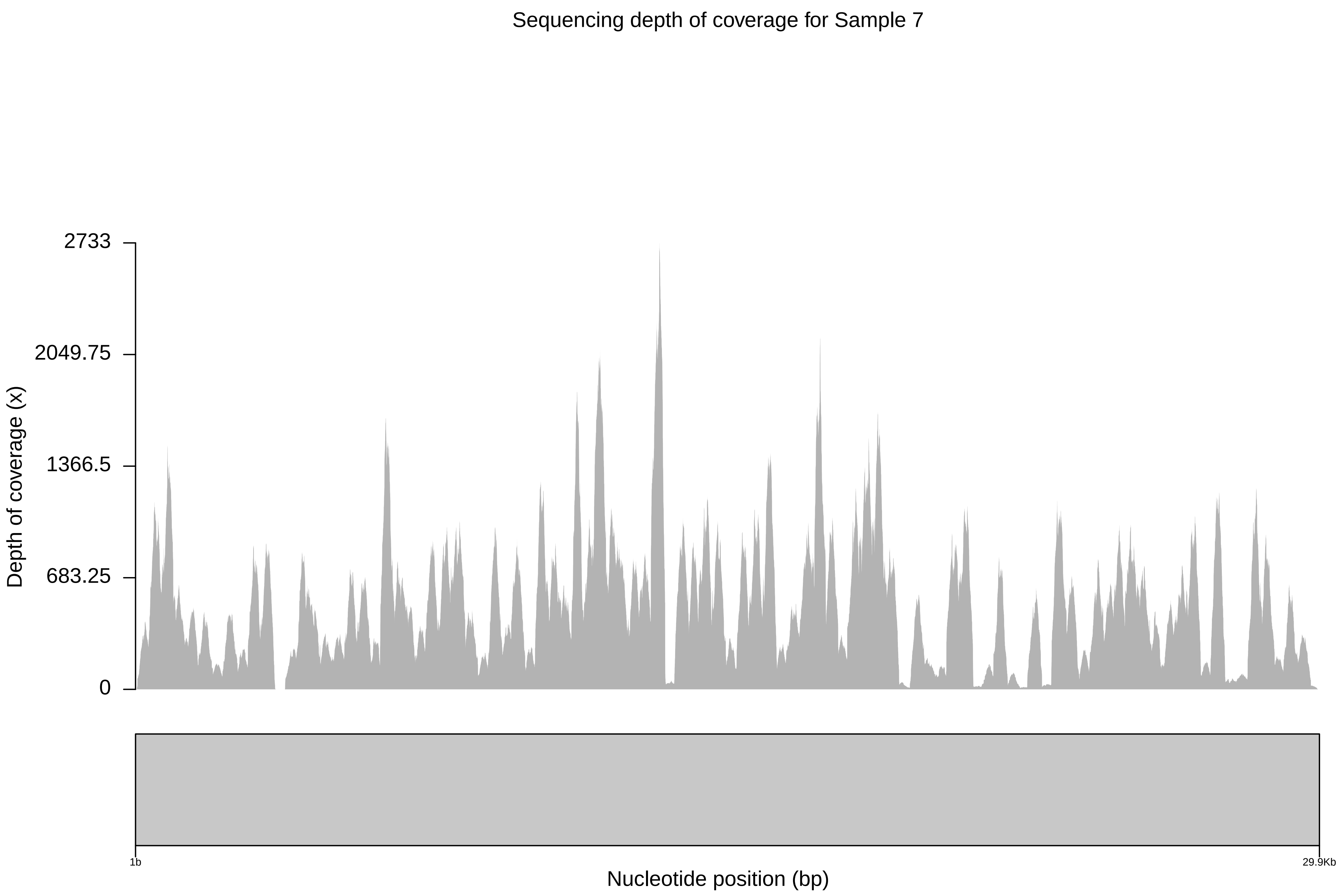

Sequencing depth of coverage for Sample 7
2733
2049.75
1366.5
683.25
0
Depth of coverage (x)
1b
29.9Kb
Nucleotide position (bp)

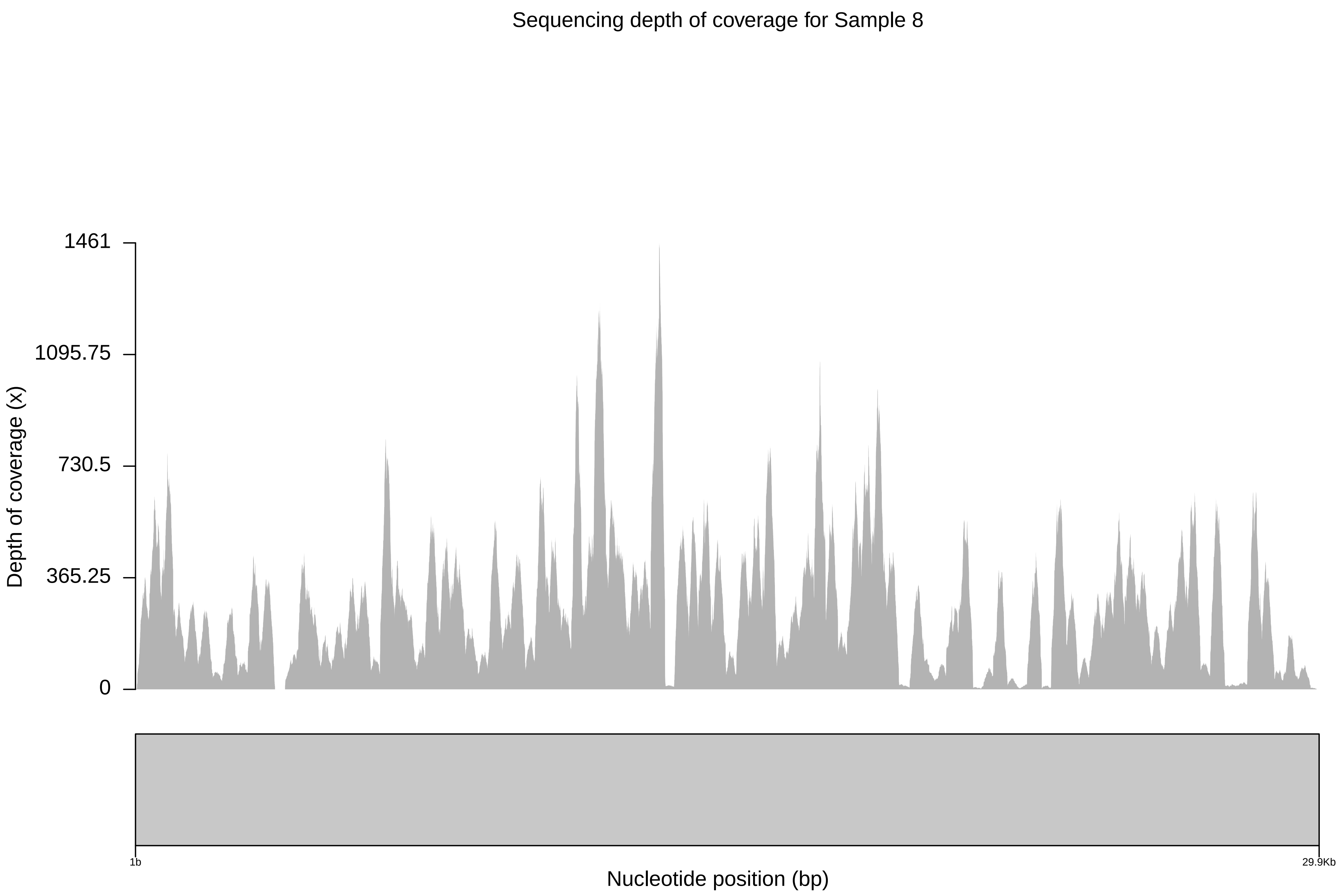

Sequencing depth of coverage for Sample 8
1461
1095.75
730.5
365.25
0
Depth of coverage (x)
1b
29.9Kb
Nucleotide position (bp)

Sequencing depth of coverage for Sample 9

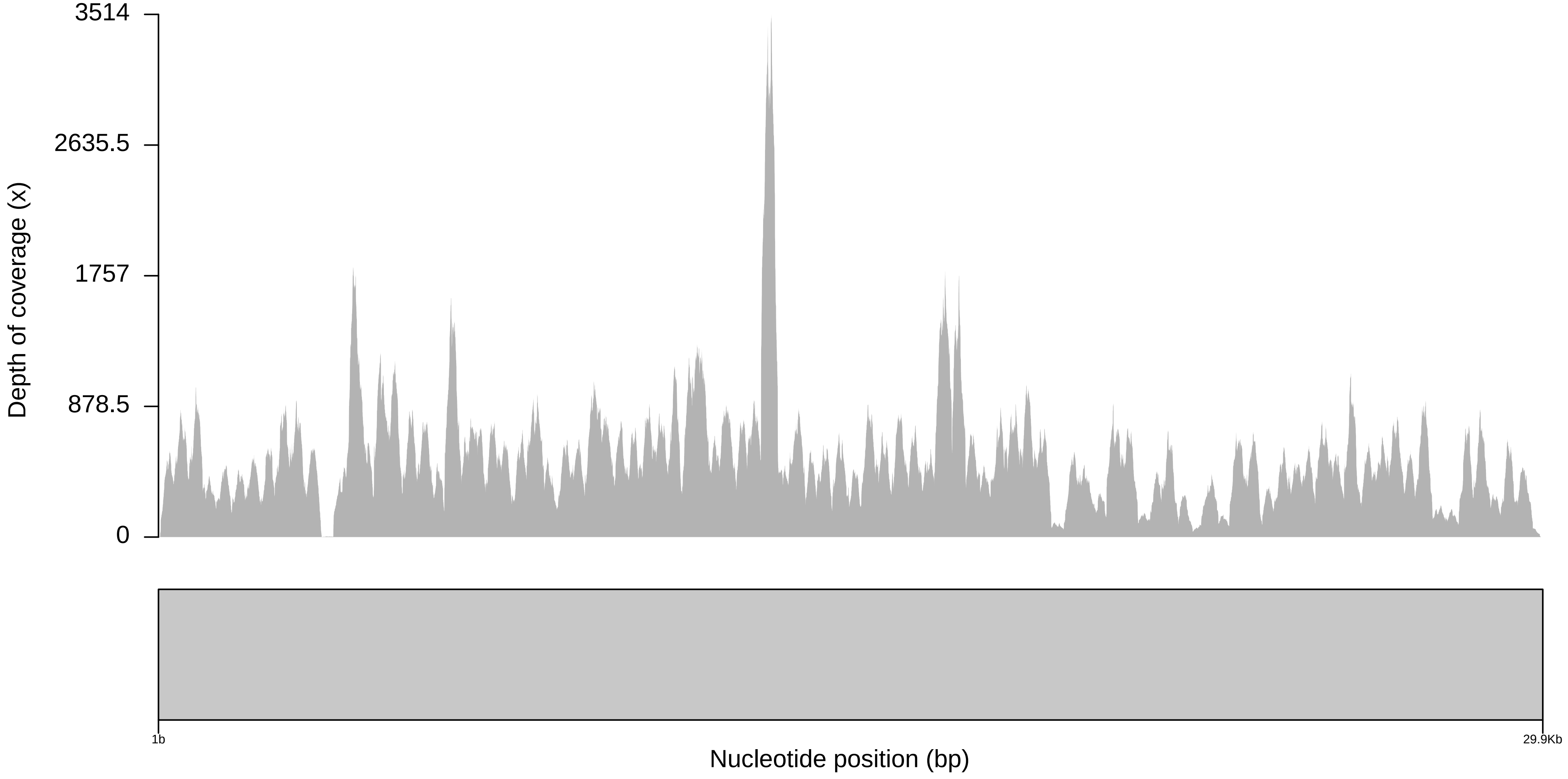

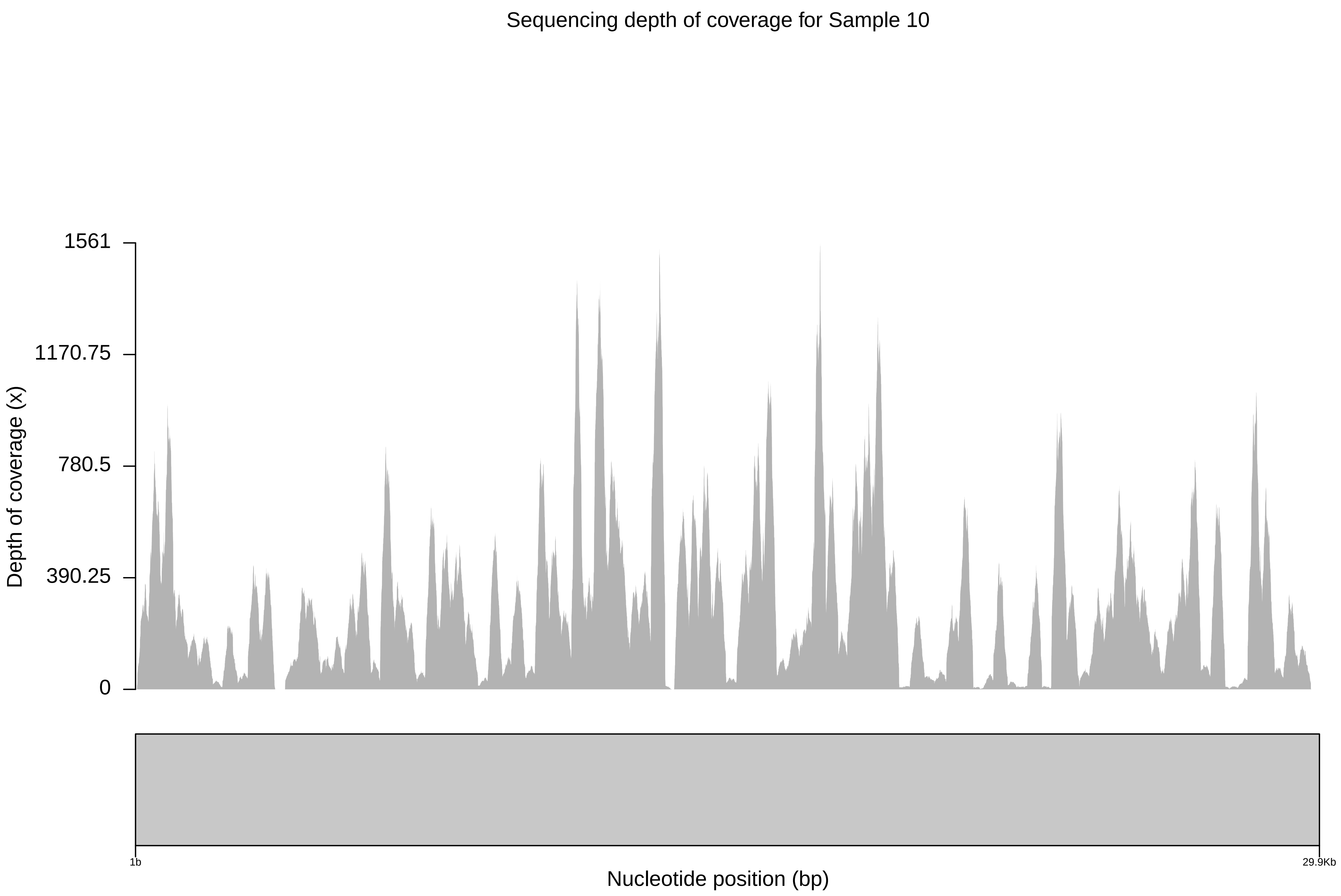
Sequencing depth of coverage for Sample 10
1561
1170.75
780.5
390.25
0
Depth of coverage (x)
1b
29.9Kb
Nucleotide position (bp)

Sequencing depth of coverage for Sample 11

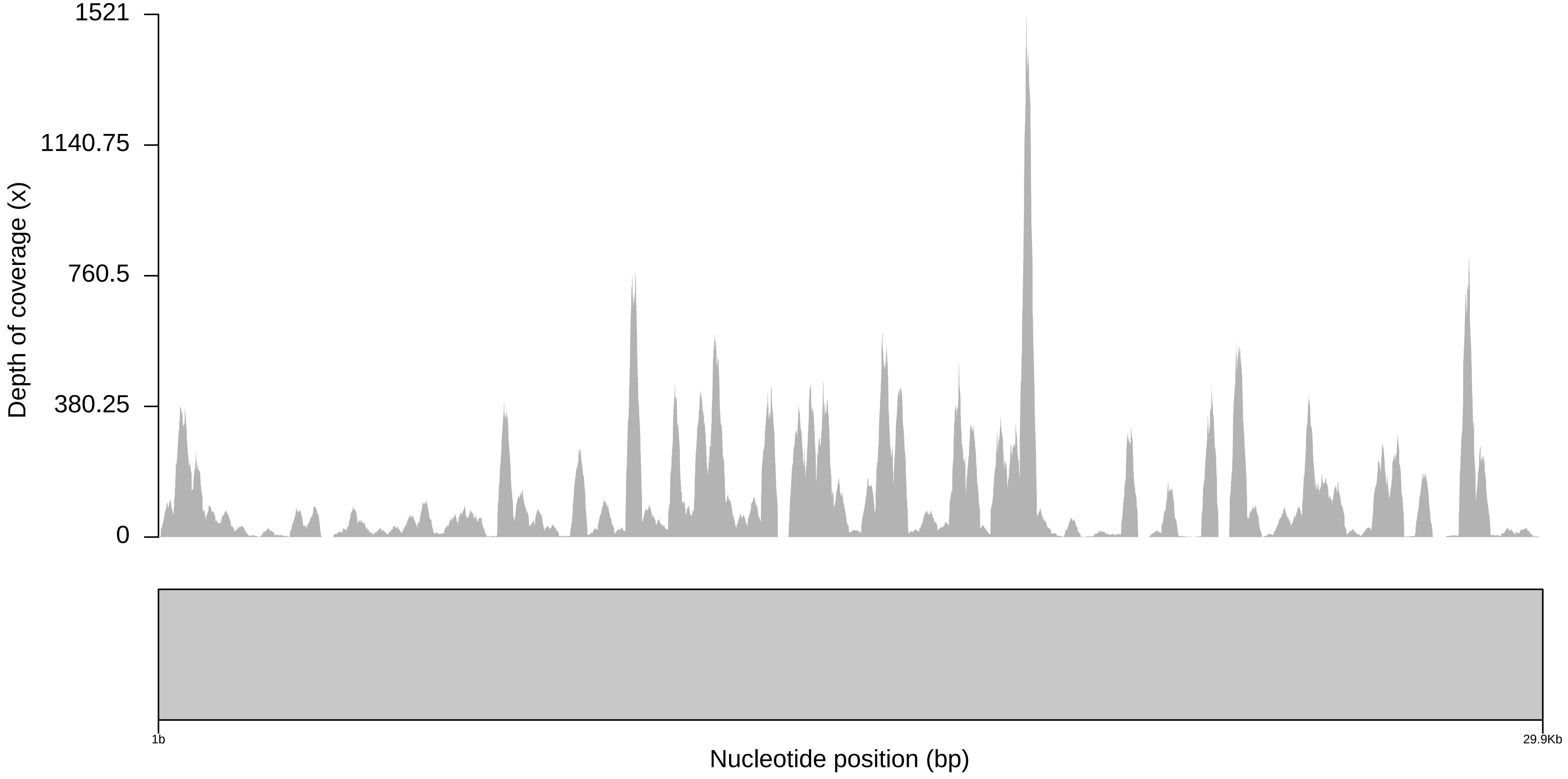

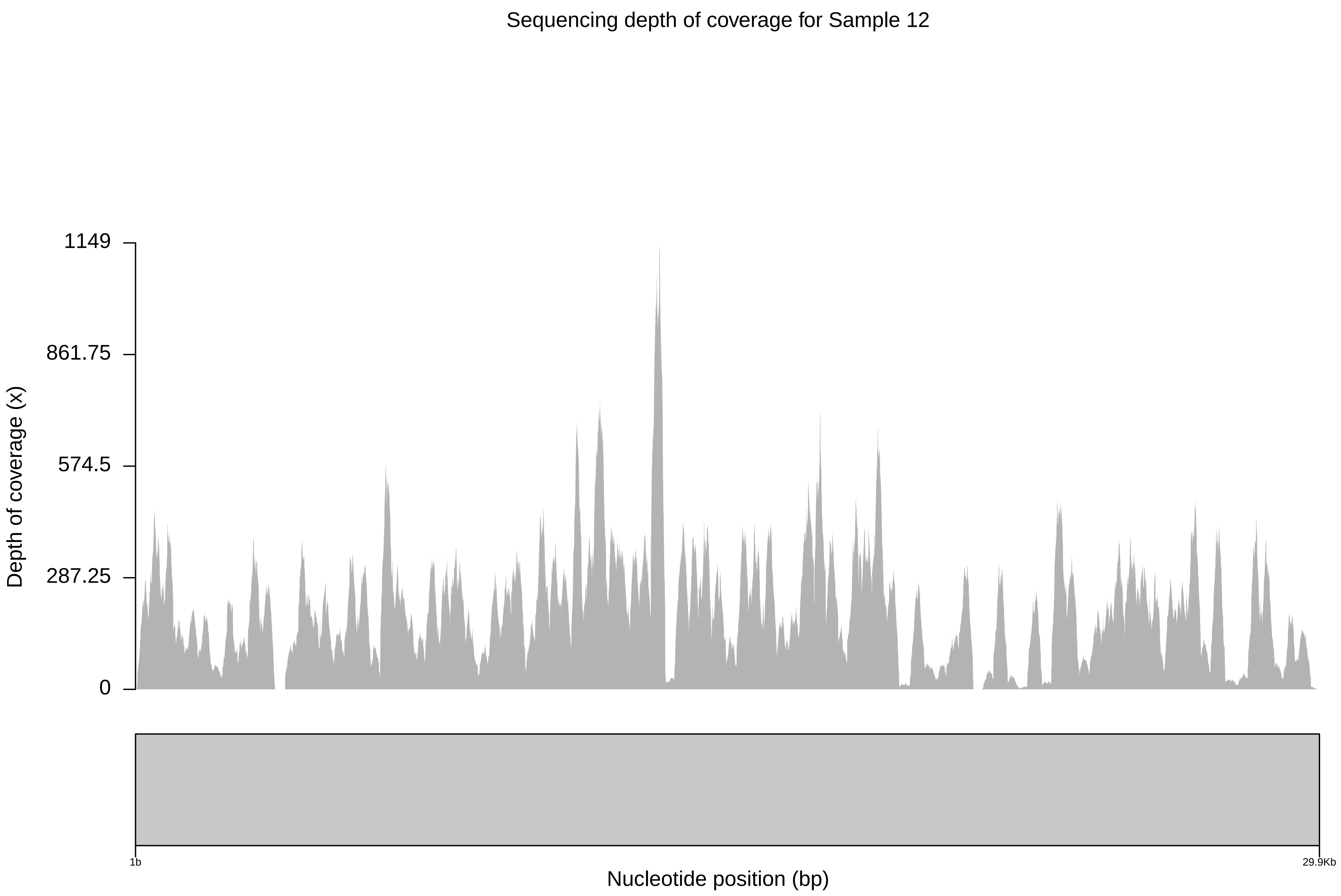

Sequencing depth of coverage for Sample 12
Depth of coverage (x)
1149
861.75
574.5
287.25
0
1b
29.9Kb
Nucleotide position (bp)
